# Development and evaluation of low-volume tests to detect and characterise antibodies to SARS-CoV-2

**DOI:** 10.1101/2022.05.03.22274395

**Authors:** Alice Halliday, Anna E Long, Holly E Baum, Amy C Thomas, Kathryn L Shelley, Elizabeth Oliver, Kapil Gupta, Ore Francis, Maia Kavanagh Williamson, Natalie di Bartolo, Matthew J Randell, Yassin Ben-Khoud, Ilana Kelland, Georgina Mortimer, Olivia Ball, Charlie Plumptre, Kyla Chandler, Ulrike Obst, Massimiliano Secchi, Lorenzo Piemonti, Vito Lampasona, Joyce Smith, Michaela Gregorova, Lea Knezevic, Jane Metz, Rachael Barr, Begonia Morales-Aza, Jennifer Oliver, Lucy Collingwood, Benjamin Hitchings, Susan Ring, Linda Wooldridge, Laura Rivino, Nicholas Timpson, Jorgen McKernon, Peter Muir, Fergus Hamilton, David Arnold, Derek N Woolfson, Anu Goenka, Andrew D. Davidson, Ashley M Toye, Imre Berger, Mick Bailey, Kathleen M Gillespie, Alistair JK Williams, Adam Finn

**Affiliations:** School of Cellular and Molecular Medicine, University of Bristol, UK; Diabetes and Metabolism, Bristol Medical School, University of Bristol, UK; School of Chemistry, University of Bristol, UK; Bristol Veterinary School, University of Bristol, UK; Population Health Sciences, Bristol Medical School, University of Bristol, UK; School of Biochemistry, Biomedical Sciences Building, University Walk, University of Bristol, UK; Diabetes Research Institute, IRCCS San Raffaele Scientific Institute, Milan, Italy; Department of Paediatric Immunology and Infectious Diseases, Bristol Royal Hospital for Children, UK; MRC Integrative Epidemiology Unit at University of Bristol, Bristol, BS8 2BN, UK; National Infection Service, UK Health Security Agency, Southmead Hospital, Bristol, UK; Academic Respiratory Unit, Bristol Medical School, University of Bristol UK; Bristol BioDesign Institute, University of Bristol, Life Sciences Building, Tyndall Avenue, Bristol BS8 1TQ, UK; Bristol Institute of Transfusion Sciences, NHS Blood and Transplant Filton, Filton, Bristol

## Abstract

Low-volume antibody assays can be used to track SARS-CoV-2 infection rates in settings where active testing for virus is limited and remote sampling is optimal. We developed 12 ELISAs detecting total or antibody isotypes to SARS-CoV-2 nucleocapsid, spike protein or its receptor binding domain (RBD), 3 anti-RBD isotype specific luciferase immunoprecipitation system (LIPS) assays and a novel Spike-RBD bridging LIPS total-antibody assay. We utilised pre-pandemic (n=984) and confirmed/suspected recent COVID-19 sera taken pre-vaccination rollout in 2020 (n=269). Assays measuring total antibody discriminated best between pre-pandemic and COVID-19 sera and were selected for diagnostic evaluation. In the blind evaluation, two of these assays (Spike Pan ELISA and Spike-RBD Bridging LIPS assay) demonstrated >97% specificity and >92% sensitivity for samples from COVID-19 patients taken >21 days post symptom onset or PCR test. These assays offered better sensitivity for the detection of COVID-19 cases than a commercial assay which requires 100-fold larger serum volumes. This study demonstrates that low-volume in-house antibody assays can provide good diagnostic performance, and highlights the importance of using well-characterised samples and controls for all stages of assay development and evaluation. These cost-effective assays may be particularly useful for seroprevalence studies in low and middle-income countries.

**Funding:** This work was supported by multiple grants to AH, AL, OF, AT, IB awarded by the Elizabeth Blackwell Institute, and funded in part by the Wellcome Trust [Grant number 204813/Z/16/Z] with additional support from Bristol Alumni and Friends. AEL is funded by a Diabetes UK/JDRF RD Lawrence Fellowship (18/0005778 and 3-APF-2018-591-A-N). The DISCOVER study was supported by donations to Southmead Hospital Charity (Registered Charity Number: 1055900). The LOGIC study was funded by The Grand Appeal (The Official Bristol Children’s Hospital Charity; a registered charity in England and Wales (1043603)) through a grant awarded to AF & AG. ACT is supported by the Wellcome Trust (217509/Z/19/Z) and UKRI through the JUNIPER consortium MR/V038613/1 and CoMMinS study MR/V028545/1. The UK Medical Research Council and Wellcome (Grant ref: 217065/Z/19/Z) and the University of Bristol provide core support for ALSPAC. NJT is a Wellcome Trust Investigator (202802/Z/16/Z), is the PI of the Avon Longitudinal Study of Parents and Children (MRC & WT 217065/Z/19/Z), is supported by the University of Bristol NIHR Biomedical Research Centre (BRC-1215-2001), the MRC Integrative Epidemiology Unit (MC_UU_00011/1) and works within the CRUK Integrative Cancer Epidemiology Programme (C18281/A29019). LIPS assay development was supported by a joint grant from Diabetes UK/JDRF (20/0006217) to KMG. IB is supported by the Wellcome Trust (106115/Z/14/Z, 221708/Z/20/Z), the ERC (contr. nrs. 834631, 963992) and the EPSRC Impact Acceleration Account EP/R511663/1. We also acknowledge funding from BBSRC/EPSRC Synthetic Biology Research Centre (BB/L01386X/1, to NDB and AMT), NHS Blood and Transplant (WP15-05, to NDB and AMT), and the NIHR Blood and Transplant Research Unit in Red Cell Products (IS-BTU-1214-10032, to NDB and AMT). This publication is the work of the authors and Alice Halliday *et al* will serve as guarantors for the contents of this paper.

**Conflict of interest:** AF is a member of the Joint Committee on Vaccination and Immunisation, the UK national immunisation technical advisory group and is chair of the WHO European regional technical advisory group of experts (ETAGE) on immunisation and *ex officio* a member of the WHO SAGE working group on COVID vaccines. He is investigator on studies and trials funded by Pfizer, Sanofi, Valneva, the Gates Foundation and the UK government. This manuscript presents independent research funded in part by the National Institute for Health Research (NIHR). The views expressed are those of the authors and not necessarily those of the NHS, the NIHR, or the Department of Health and Social Care.

## Introduction

The COVID-19 pandemic has caused over 6 million deaths and significant morbidity worldwide and major disruption to many societies (1). Mass testing for antibodies specific for SARS-CoV-2 plays an important role in understanding prevalence and transmission at the population level and has become a pillar of COVID-19 surveillance in many countries including the UK (2). In contrast to tests detecting the virus, antibody assays can confirm previous infection and are particularly useful for identifying undetected, often asymptomatic cases (3). This provides a more sensitive and practical approach to estimating prevalence than repeated testing of symptomatic individuals. Moreover, measuring antibody responses to different antigens and detecting specific antibody isotypes provides insights into levels of immunity in populations and mechanisms of immune-protection. Over time SARS-CoV-2 antibody testing is playing an increasing role in shaping vaccine evaluation and policy (4, 5). In low and middle income countries, less viral antigen/PCR testing is performed, thus data on infection and immunity gathered through simple and affordable tests is essential to guide vaccination rollout (6).

While many approaches to measuring antibodies to SARS-CoV-2 have been reported previously (7, 8), there is considerable variability in reported seropositivity rates determined using different assays/platforms, so that robust conclusions relating to population level exposure and immunity are elusive. Lab-based commercial assays have been widely evaluated in target populations in high income settings, offer good diagnostic performance, and have been deployed effectively to determine serological antibody status as well as to support clinical diagnoses (9). However, these assays are costly, often require dedicated specialist equipment, large sample volumes, and in general do not fully characterise the antibody response in terms of isotype. In contrast, lateral flow antibody assays require very low sample volumes and can be cheaper. These user-facing tests are suitable for large population surveillance and surveys, but have shown sub-optimal performance for detection of SARS-CoV-2 specific antibodies in evaluation studies (10). Sub-optimal performance of antibody assays can lead to uncertainty around persistence of antibodies and associated immune protection. Non-commercial (‘in-house’) assays have played an essential role in characterising the antibody responses in various large cohort studies (11, 12), but many lack standardization and have been optimised/evaluated using relatively small numbers of samples making their performance unclear (13). A range of high-performance platforms such as ELISA and Luciferase Immunoprecipitation System (LIPS) assays have been reported and widely used for the measurement of SARS-CoV-2 antibodies (7, 14). Although less well known than ELISA, the LIPS platform offers an alternative method for measuring SARS-CoV-2 specific antibody responses that is more amenable to the use of competition (selecting for high affinity antibodies), and has potential to improve specificity for target antigens/epitopes (14, 15). LIPS also offers a wider dynamic range, allowing quantification of antibody responses using low sample volumes.

We therefore sought to utilise a varied and well characterised set of samples from recent COVID-19 cases (known positives) and pre-pandemic (known negative) individuals to develop and retrospectively evaluate in-house antibody assays for detecting recent infection. Multiple studies have reported differing antibody titres and profiles in groups with varying clinical outcomes (12, 16), as well as varying levels of cross-reactive responses among pre-pandemic samples from individuals of different ages (17). The COVID-19 cases therefore included severe and mild hospitalised PCR-confirmed and clinically suspected cases as well as PCR-confirmed mild and asymptomatic SARS-CoV-2 community infections (18) using samples collected during the early stages of the pandemic (2020). These were combined with large numbers of pre-pandemic sera from multiple collections (known negatives) and distributed across distinct stages of assay development and evaluation. The assays included total antibody and isotype-specific, standardised, low serum volume assays on two different platforms (ELISA and LIPS) and a novel LIPS bridging format with high throughput potential. To facilitate interpretation of the assays, we also sought to relate the results of our binding antibody assays to functional responses by comparing with two different neutralisation assay platforms. Finally, we describe the process of reporting binding antibody in international units and demonstrate the deployment of the assays in a population of healthcare workers with unknown serological and infection status.

## Results

### Sample collections and study flow for assay evaluation

We collected well characterised plasma and serum samples from multiple sources and distributed them into sets for distinct stages of assay development and evaluation (Figure 1). The ‘known negative’ samples were pre-pandemic samples from a range of donor types including adult blood donors, and adults and children involved in research studies. A small group of adult hospitalised patients with pneumonia/other pleural conditions were also included (Figure 1A). The cohort of COVID-19 cases were recruited via two routes in 2020 (prior to vaccine rollout): 1) convalescent hospital workers identified after receiving a positive PCR test, and 2) patients confirmed and/or clinically suspected to have COVID-19 at the point of recruitment during inpatient/outpatient hospital/secondary care visits in Bristol, UK (Figure 1B). The COVID-19 cohort included the full spectrum of disease (asymptomatic to severe/fatal) and encompassed a range of times since symptom onset or PCR confirmation (those without PCR confirmation were clinically suspected based on symptom presentation in hospital). The sample sets were used for development and evaluation as shown in Figure 1C, with only the optimal screening assays being included in the full diagnostic evaluation process.

**Figure 1.**
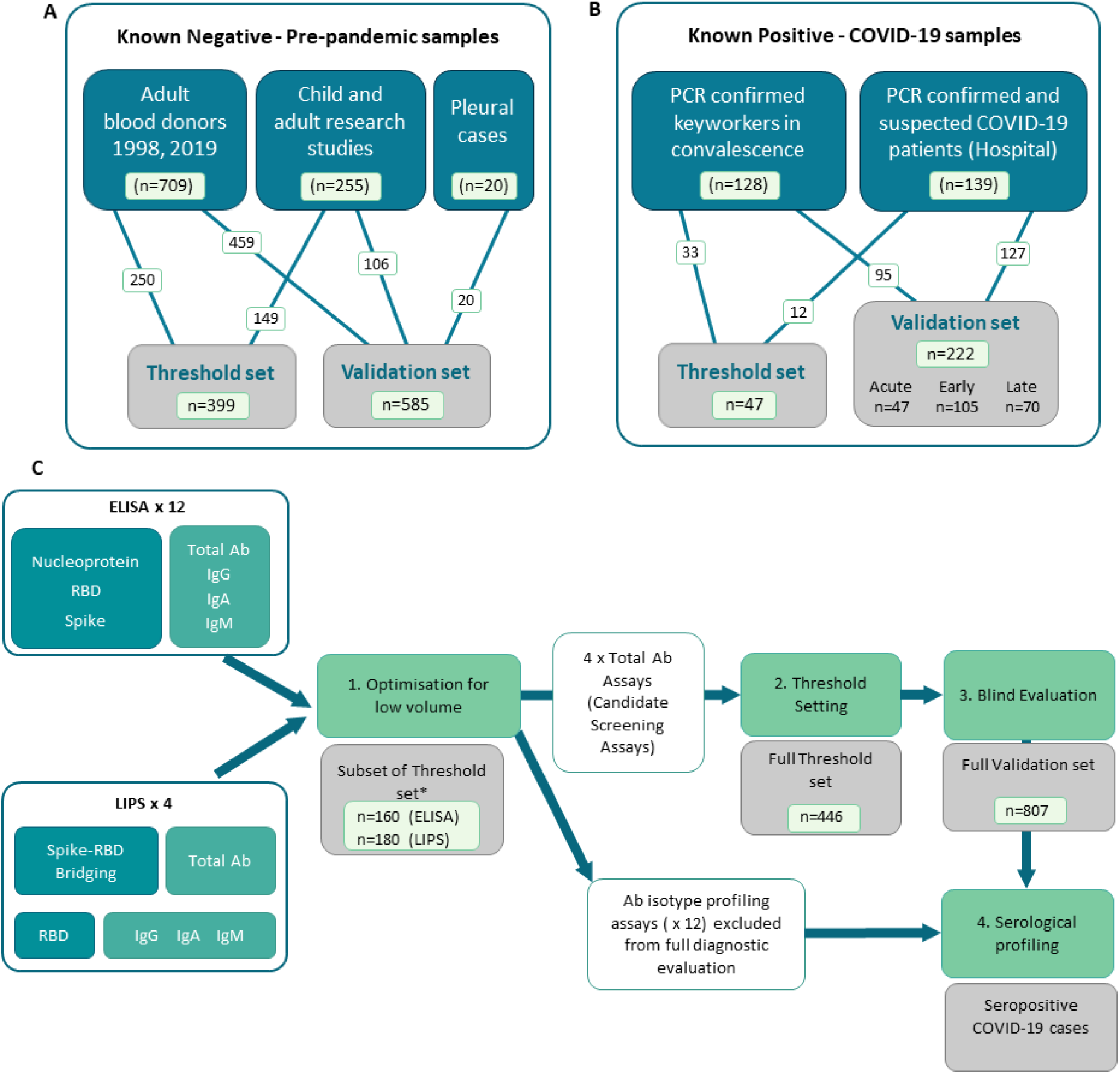
Distribution of samples into sets, and flow diagram of assay development and evaluation. In-house COVID-19 antibody assays using pooled controls and a set of well characterised samples to first optimise and then set thresholds for positive and negative. A) Known negative samples were all collected pre-pandemic (i.e. pre-December 2019) from various Biobanks and included adult and child plasma and serum samples from research studies, adult blood donors and a small collection of samples from hospitalised cases of pleurisy. B) Known positive samples were collected from a full spectrum of COVID-19 adult cases, including RT-PCR confirmed COVID-19 cases recruited in convalescence from the community and PCR confirmed and clinically suspected COVID-19 cases recruited in hospitals. C) Flow diagram showing assay development, selection of screening assays and diagnostic evaluation. A total of 16 assays including isotype-specific and total antibody assays on both ELISA and LIPS platforms were optimised using a subset of the threshold set of these samples (n=160 or n=180depending on assay platform) for initial selection of candidate screening assays (Stage 1). * Note that whilst the number of threshold set samples used for optimisation was comparable for ELISA and LIPS assays, the exact samples used differed slightly due to low samples volume for use across 16 assays. The assays which performed best for each antigen/platform combination were then taken forward for the full threshold setting to determine optimal thresholds for specificity (stage 2) including the remaining n=∼300 samples for all 4 screening assays (i.e. total of n=446). Candidate screening assays with pre-defined thresholds were then deployed on a blind validation cohort containing n=222 samples from COVID-19 cases and n=585 pre-pandemic samples. The performance characteristics were defined using the validation set to guide utility of deployment. The 12 x non-screening assays were subsequently used only for profiling of seropositive samples (stage 4) and comparing to functional assays.

### Assay optimisation and selection of screening assays

By building on an ELISA protocol made widely available at the start of the pandemic, we used a subset of the threshold set of samples (n=160) to optimise assay conditions to measure antibodies specific to three antigens from SARS-CoV-2: the Nucleocapsid (N), the receptor binding domain (RBD) of Spike, and the stabilised trimeric Spike protein, with a focus on achieving discrimination between pre-pandemic and COVID-19 samples using small serum/plasma volumes (<10 µl). For each antigen, a suite of ELISAs was set up to measure either total antibody/Pan Immunoglobulin using a commercially available ‘anti human IgG’ secondary antiserum which detects the Fab region (i.e., anti-IgG H+L) and therefore detects all isotypes, (hereafter labelled ‘Pan’), or IgG, IgA and IgM isotypes (using class-specific secondary antibodies) (Figure S1). We generated a pooled serum ‘standard’ from donors with high responses to all antigens that was used to standardise between plates (Figure S2A). The optimal dilutions for the standards, controls and samples to achieve discrimination between pre-pandemic and COVID-19 samples were determined. We found that a single sample dilution, normalised to a top standard control, could provide improved or equivalent discrimination between groups when compared to using AUC calculated from a 4-point dilution series or deriving an interpolated value (respectively) (Figure S2B-C). Since the aim was to optimise assays for discrimination, but also low sample-volume and high throughput, we elected to report ELISA results as Normalised ODs which are the simplest to perform. For each antigen, the Pan (total antibody) ELISAs outperformed the isotype specific assays in discriminating between pre-pandemic samples and those from COVID-19 cases, as demonstrated by greater AUCs after receiver operator characteristic (ROC) analysis (Figure S2A).

A suite of LIPS assays measuring antibodies specific to the RBD, were developed based on protocols outlined in (14), a study in which detection of serum RBD specific antibodies was found to be associated with survival in hospitalised cases of COVID-19. For measuring IgG and IgA isotypes, using unlabelled RBD in competition with Nluc-labelled RBD was found to improve discrimination between pre-pandemic and COVID-19 cases (Figure S3A&B) whereas no optimal level of competition was identified for IgM. For detection of total antibody with high affinity for RBD, the novel Spike-RBD bridging assay format was adopted in which plates coated with trimeric stabilised Spike could bind samples prelabelled with Nluc-RBD (Figure S1B). All LIPS-based assay results are reported in interpolated units using the internal pooled serum standard. As with the Pan ELISAs, the Spike-RBD bridging assay demonstrated superior AUCs after ROC analysis amongst the LIPS assays for discrimination between COVID-19 cases and pre-pandemic controls when ROC curve analysis was performed (Figure 2C). Despite using the same target antigen, the RBD LIPS assays provided higher AUCs than the RBD-specific ELISA assays for all antibody isotypes when the same samples sets were compared (Figure S3C).

**Figure 2.**
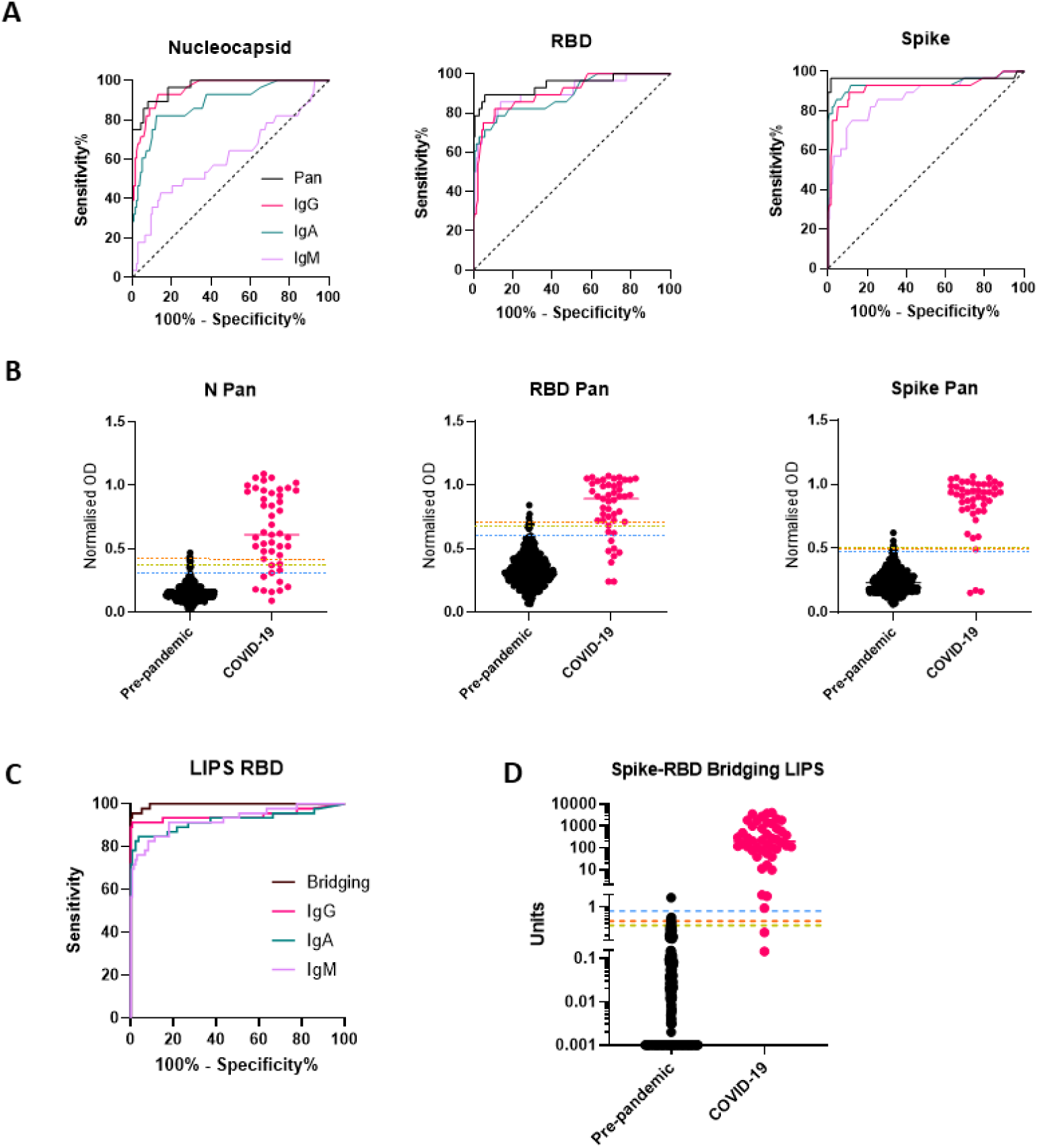
Selection of screening assays and threshold setting with the threshold set. A subset of the threshold set samples were used for assay optimisation and then to compare performance of isotype/total antibody specific assays for each antigen/platform combination. A) ROC curves showing the relative performance of ELISAs using different secondary antibodies (Pan total antibody (Black); IgG (pink); IgA (green), IgM (purple) in a cohort of n=27 COVID-19 samples and n=133 pre-pandemic samples from the threshold set. For all three antigens, the Pan/total antibody assays provided the best performance as evidenced by highest AUCs. B) Scatterplots showing the individual Normalised OD readings for all three ELISA screening assays (N Pan, RBD Pan and Spike Pan) in the full threshold set of n=45 COVID-19 samples and n=399 pre-pandemic), with the median represented by a line for each group and the three thresholds for each assay indicated with a line across the plot: 1 – the 99^th^ percentile of pre-pandemic levels (orange dashed line); 2 - The 98^th^ percentile (yellow dashed line); 3 – Youden’s index (blue dashed line). C) ROC curves for all LIPS assays deployed on a subset of the threshold set (n= 46 COVID-19 cases and n=134 pre-pandemic) showing optimal performance with the Spike-RBD Bridging LIPS assay, which was therefore taking forward for full threshold setting on the full threshold set (n=446) with results shown in the scatterplot in D) (interpolated unit values shown on y axis with log10 scale and broken axis to allow visualisation of thresholds. To ensure all results were plotted, a result of zero units was assigned a value of 0.001 for this graph. The three thresholds are indicated: 1 – the 99^th^ percentile of pre-pandemic levels (orange dashed line); 2 - The 98^th^ percentile (yellow dashed line); 3 – Youden’s index (blue dashed line).

In summary, we developed 16 antibody assays for detection of COVID-19 antibody responses using <5 µl serum samples in a single dilution, which can be scaled to high throughput. Since the total antibody assays (i.e., N, RBD and Spike Pan ELISAs and the Spike-RBD bridging LIPS) performed best for discrimination in the optimisation stage, these were designated as the best candidates for screening assays to identify COVID-19 seropositive individuals.

### Threshold setting

Candidate screening assays (n=4) were run on samples from the full threshold set consisting of 399 pre-pandemic and 47 COVID-19 case samples (Table S1); the predominance of pre-pandemic samples in this cohort highlights the aim to achieve optimal specificity. Thresholds were determined using three distinct criteria: 1) The 99^th^ percentile of the negative (pre-pandemic) controls; 2) The 98^th^ percentile of the negative (pre-pandemic) controls; 3) The highest Youden Index to achieve balanced sensitivity and specificity (Figure 2B-C) and assay performance in the threshold set was determined. In this sample set, all assays provided good discrimination between COVID-19 and pre-pandemic samples (AUCs ranging from 0.95-0.997), with the Spike-RBD bridging LIPS assay providing optimal and almost perfect performance (AUC = 0.997, 95% CI 0.993-1.001) (Table S2). We did not observe differences in pre-pandemic total antibody assay signals between adults, teens and children (Figure S4A). We compared background signal levels in serum and plasma and found no statistical differences in signal between matched plasma/serum from pre-pandemic donors (Figure S4). Thus, using both serum and plasma samples, thresholds for each assay were set which were identical for all age groups.

### Blind evaluation of screening tests to detect confirmed and suspected COVID-19 cases

The 4 screening assays measuring total antibodies were tested on the full validation sample set (n=807) in a blinded fashion. To explore sensitivity of candidate assays for different periods post infection, the COVID-19 samples were divided into 3 time periods for diagnostic evaluation such that no repeat samples were included in each group: Acute (< 21 days post symptom onset (p.s.o) or PCR test), Early Convalescent (3-12 weeks p.s.o), Late Convalescent (>12 weeks p.s.o) (Table 1). Results for the 4 screening assays are presented either as dot plots or ROC curves (Figure 3A, C, D, F). Sensitivity and specificity estimates (and 95% CI) of the assays at each pre-defined threshold are reported (Table 2) and presented in (Figure 3B, E). High specificity for recent COVID-19 infection was maintained from the threshold setting set, with all thresholds providing >96% specificity (and most >98%); for each assay, the threshold providing at least 98% specificity whilst achieving optimal sensitivity was selected (Table 2), except for the Spike Pan ELISA where the highest specificity provided was 97.3% (95% CI 95.9-98.5). Of note, the optimal threshold method differed across the 4 assays, e.g. for the N Pan ELISA the 98^th^ percentile performed optimally, whilst for the Spike-RBD bridging LIPS it was the Jmax value (highest Youden’s index). The screening assays generally displayed low levels of intra- and inter-assay variation; coefficients of variation were between 1.8% and 23.4% for QC samples in the positive range (Table S3).

**Figure 3.**
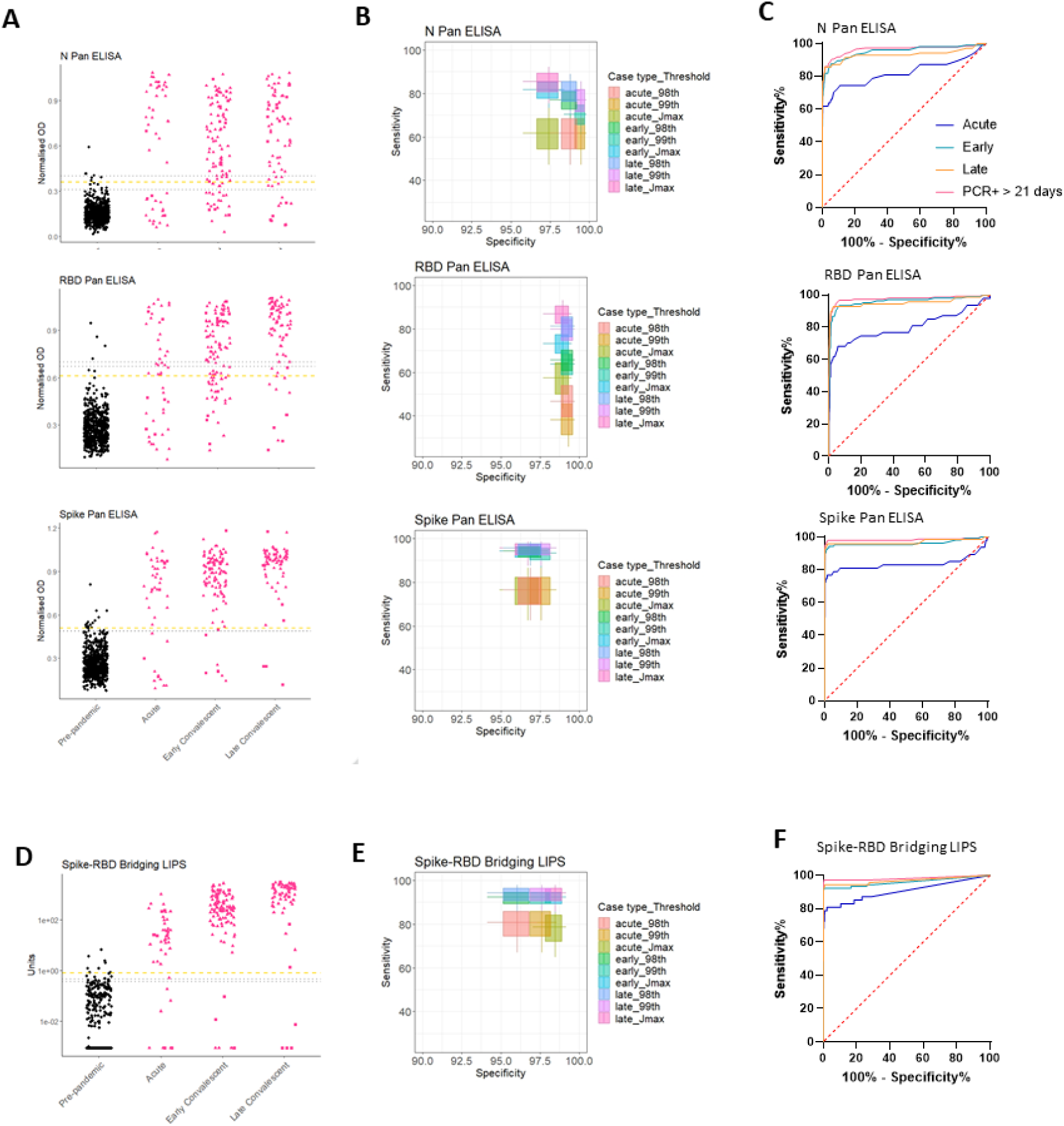
Performance of screening assays in validation cohort. Dot plots showing assay results for the 3 screening ELISAs (N, RBD and Spike Pan (A)) and the Spike-RBD Bridging LIPS (D); black dots represent pre-pandemic samples and pink dots represent COVID-19 samples separated by different periods post infection: Acute; Early Convalescent; Late Convalescent. One misrepresentative highly positive pre-pandemic result has been removed from (D) as analysis after unblinding indicated it was the result of human error after blinding (it still is included in the ROC/sensitivity analyses). (B, E) Boxplots indicating sensitivity and specificities for COVID-19 performed by each assay at each of the pre-defined thresholds. (C,F) ROC curves indicating performance of each assay for differentiating acute (< 21 days p.s.o.; blue), early convalescent (21 days – 12 weeks p.s.o; turquoise) or late convalescent (> 12 weeks p.s.o.; orange) COVID-19 cases from pre-pandemic samples in the blind validation cohort. The performance for detection of the most likely to be ‘true seropositive’ COVID-19 cases is also included, i.e. those who were sampled after 21 days after a confirmed PCR test (pink).

**Table 1.**
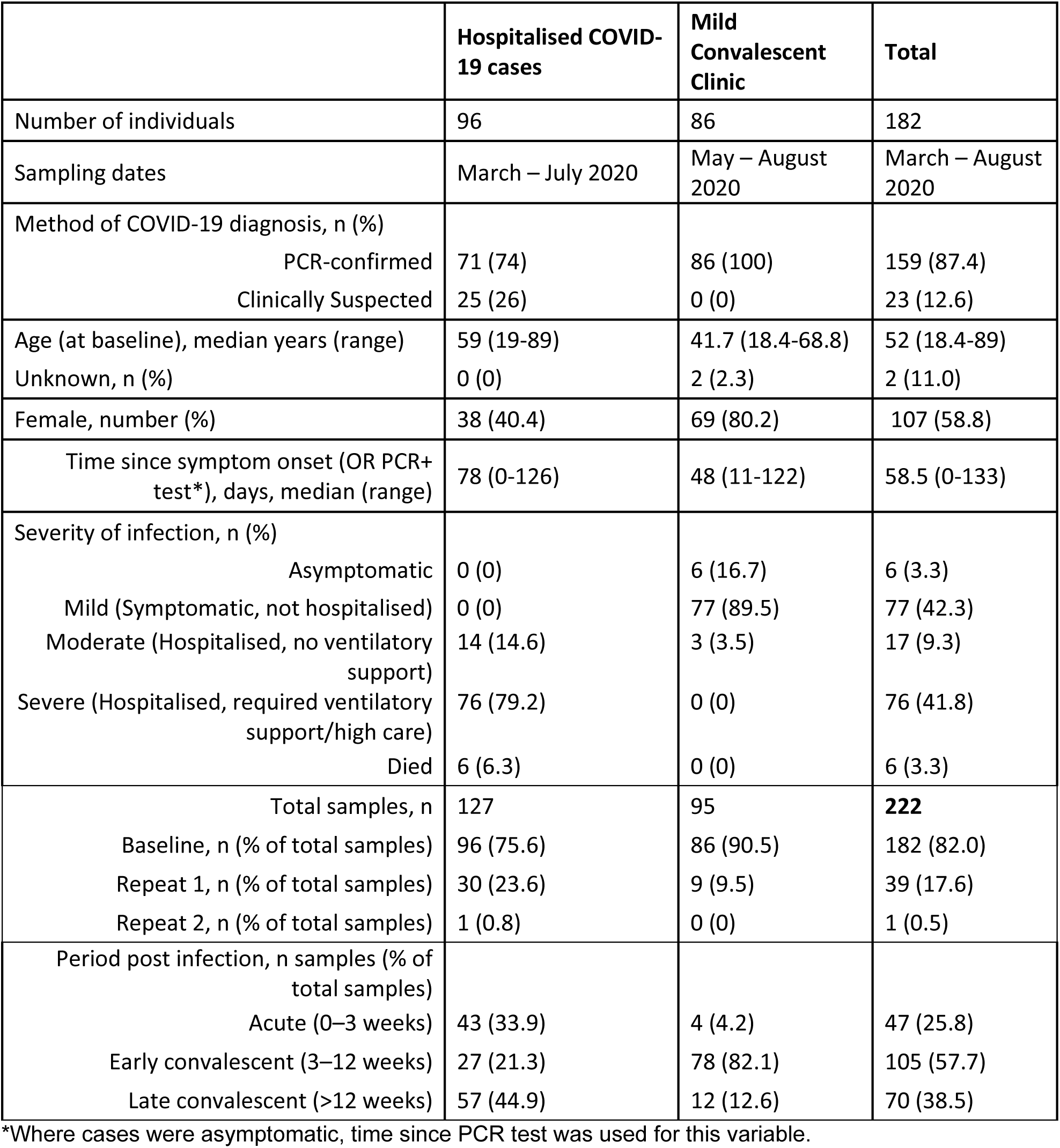
COVID-19 cases in validation cohort. Clinical and demographic features relating to the n=182 individuals diagnosed with COVID-19 whose samples (n=222) were included to assess sensitivity of the screening assays. Individuals were recruited via two main routes – either as part of secondary care in hospital, or community cases invited to a convalescent clinic at least 14 days after symptom onset. Percentages are from total of individuals unless otherwise stated.

|  | **Hospitalised COVID-19 cases** | **Mild Convalescent Clinic** | **Total** |
| --- | --- | --- | --- |
| Number of individuals | 96 | 86 | 182 |
| Sampling dates | March – July 2020 | May – August 2020 | March – August 2020 |
| Method of COVID-19 diagnosis, n (%) |  |  |  |
| PCR-confirmed | 71 (74) | 86 (100) | 159 (87.4) |
| Clinically Suspected | 25 (26) | 0 (0) | 23 (12.6) |
| Age (at baseline), median years (range) | 59 (19-89) | 41.7 (18.4-68.8) | 52 (18.4-89) |
| Unknown, n (%) | 0 (0) | 2 (2.3) | 2 (11.0) |
| Female, number (%) | 38 (40.4) | 69 (80.2) | 107 (58.8) |
| Time since symptom onset (OR PCR+ test*), days, median (range) | 78 (0-126) | 48 (11-122) | 58.5 (0-133) |
| Severity of infection, n (%) |  |  |  |
| Asymptomatic | 0 (0) | 6 (16.7) | 6 (3.3) |
| Mild (Symptomatic, not hospitalised) | 0 (0) | 77 (89.5) | 77 (42.3) |
| Moderate (Hospitalised, no ventilatory support) | 14 (14.6) | 3 (3.5) | 17 (9.3) |
| Severe (Hospitalised, required ventilatory support/high care) | 76 (79.2) | 0 (0) | 76 (41.8) |
| Died | 6 (6.3) | 0 (0) | 6 (3.3) |
| Total samples, n | 127 | 95 | **222** |
| Baseline, n (% of total samples) | 96 (75.6) | 86 (90.5) | 182 (82.0) |
| Repeat 1, n (% of total samples) | 30 (23.6) | 9 (9.5) | 39 (17.6) |
| Repeat 2, n (% of total samples) | 1 (0.8) | 0 (0) | 1 (0.5) |
| Period post infection, n samples (% of total samples) |  |  |  |
| Acute (0–3 weeks) | 43 (33.9) | 4 (4.2) | 47 (25.8) |
| Early convalescent (3–12 weeks) | 27 (21.3) | 78 (82.1) | 105 (57.7) |
| Late convalescent (>12 weeks) | 57 (44.9) | 12 (12.6) | 70 (38.5) |

\*Where cases were asymptomatic, time since PCR test was used for this variable.

**Table 2.** Performance of total antibody screening assays in blind diagnostic evaluation using the validation set (n=807 samples). For each candidate screening assays, one value for specificity of the assays is reported, and sensitivity estimates are provided for COVID-19 samples taken at different time periods (Acute, Early Convalescent and Late Convalescent) post infection are reported for each of the threshold methods. Confidence intervals (95% CI) are given for each estimate. The optimal threshold methods was determined by finding the value which provided at least (or as near to) 98% specificity, whilst offering the optimal sensitivity.

|  | Specificity at defined thresholds (95% CI) |  |  | Sensitivity for Acute COVID-19 at defined thresholds (95% CI) |  |  | Sensitivity for Early Convalescent COVID-19 at defined thresholds (95% CI) |  |  | Sensitivity for Late COVID-19 at defined thresholds (95% CI) |  |  | Optimal Threshold Value (method) |
| --- | --- | --- | --- | --- | --- | --- | --- | --- | --- | --- | --- | --- | --- |
| Threshold* | 1 | 2 | 3 | 1 | 2 | 3 | 1 | 2 | 3 | 1 | 2 | 3 |  |
| N Pan ELISA | 99.5<br>(98.4 - 99.8) | 98.8<br>(97.5 - 99.4) | 97.4<br>(95.8 - 98.4) | 61.7<br>(47.2 - 74.4) |  |  | 70.5<br>(61.1 - 78.4) | 77.1<br>(68.1 - 84.2) | 81.9<br>(73.3 - 88.2) | 77.1<br>(65.9 - 85.5) | 81.4<br>(70.6 - 88.9) | 85.7<br>(75.4 - 92.1) | (2) |
| RBD Pan ELISA | 99.3<br>(98.2 - 99.7) |  | 99<br>(97.7 - 99.5) | 38.3<br>(25.6 - 52.8) | 46.8<br>(33.1 – 61) | 57.4<br>(43.1 - 70.7) | 63.8<br>(54.2 - 72.4) | 65.7<br>(56.1 - 74.2) | 73.3<br>(64.1 - 80.9) | 80<br>(69 - 87.8) | 81.4<br>(70.6 - 88.9) | 87.1<br>(77.1 - 93.2) | (3) |
| Spike Pan ELISA | 97.3 (95.9 - 98.5) | 96.7<br>(95.2 – 98.1) | 96.7<br>(95.2 – 98.1) | 76.6<br>(62.5 - 86.6) |  |  | 93.3<br>(88.6 – 97.1) | 94.3<br>(87.8 - 98.1) |  | 95.7<br>(87.5 - 98.6) |  |  | (1) |
| Spike-RBD Bridging LIPS | 97.6<br>(96 - 98.6) | 96<br>(94.1 - 97.4) | 98.5<br>(97.1 - 99.2) | 80.9<br>(67.1 - 89.7) | 80.9<br>(67.1 - 89.7) | 78.7<br>(64.8 - 88.2) | 92.4<br>(85.5 - 96.1) |  |  | 94.3<br>(85.7 - 97.8) |  |  | (3) |

\*Threshold method key: 1 - 99<sup>th</sup> Percentile; 2 - 98<sup>th</sup> percentile; 3 - Jmax i.e., highest Youden Index

We performed a sensitivity analysis to evaluate whether inclusion of samples from clinically suspected COVID-19 cases, who were not virologically confirmed, and/or those samples collected within 21 days p.s.o (i.e. in the Acute group), was detrimental to diagnostic performance when compared to including only samples from RT-PCR confirmed individuals 3 weeks p.s.o (one sample per donor only) (Figure 3C, F, Table S4). The performance of each assay was best in the RT-PCR confirmed group, suggesting that some of the suspected COVID-19 cases may not have been infected with SARS-CoV-2.

The best performing assays for all groups were the Spike Pan ELISA and the Spike-RBD bridging LIPS assay which provided comparable sensitivities for detection of COVID-19 cases (i.e. between 92.4-95.7% sensitivity for COVID-19 cases >21 days p.s.o, depending on the group of interest (Table 2, S4, Figure 3), and good specificity (between 96.7-98.5% depending on the threshold used).

### The Spike-RBD Bridging LIPS assay and Spike Pan ELISA perform better than a commercial high-volume assay for detection of recent COVID-19 cases

We compared the sensitivity of the commercially available Roche Elecsys (high volume) serum assay to the ELISA and LIPS screening assays using samples from 184 COVID-19 cases from the validation for which sufficient volume of sample was available. The Spike-RBD Bridging LIPS assay and the Spike Pan ELISA provided optimal sensitivity for COVID-19 cases, with both providing 93% sensitivity for RT-PCR confirmed cases whilst the Roche assay provided 85.5% (Figure 4A and Table S5); the N Pan and RBD Pan ELISAs provided 76.97 and 76.32% sensitivity respectively. Within the PCR confirmed group of patients, the Spike-RBD Bridging LIPS and Spike Pan ELISA assays detected 21 of the 22 samples that were found negative with the Roche assay despite requiring >100 x less sample volume.

**Figure 4.**
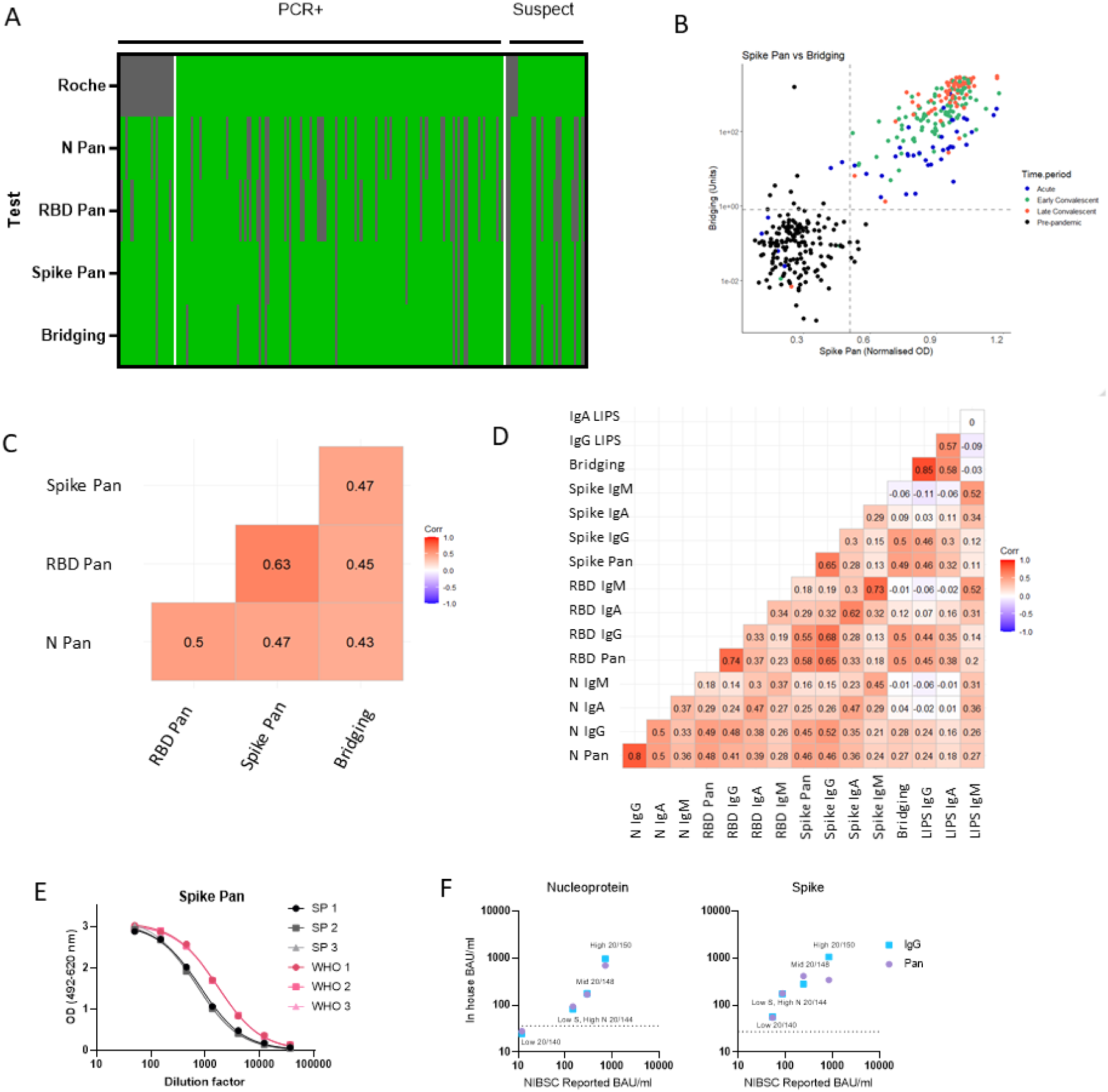
Comparing sensitivity of screening assays to a commercial assay (Roche Elecsys nucleocapsid) and correlation/concordance of all antibody assay results in samples within the validation set. (A) Heatmap comparing sensitivity of the 4 screening assays to the commercially available Roche Elecsys nucleocapsid assay in a cohort n=184 individuals with RT-PCR confirmed or suspected COVID-19 (from the validation set). Green indicates above positive threshold, grey negative. B) Scatterplot showing the quantitative readouts from the top performing screening assays – Spike Pan ELISA and Spike-RBD Bridging LIPS assay with their best performing threshold indicated with dashed lines. C) Correlogram reporting correlation coefficients using Kendall’s tau of all 4 screening assays in full validation set including pre-pandemic samples (n=806). (D) Correlogram showing the relationship between results of all 16 antibody assays on COVID-19 cases (n=222).

A comparison of the Spike Pan ELISA and Spike-RBD bridging LIPS assays revealed very good concordance for the detection of COVID-19 cases, but less so for the responses recorded from pre-pandemic samples, where different individuals were found to be positive on each assay (Figure 5B). Kendall’s tau correlations were performed to compare screening assay results (Normalised ODs) on the full validation set. Strongest agreement was observed between the Spike and RBD Pan ELISA responses (Figure 5C). All isotype specific assays were deployed on the seropositive COVID-19 cases; within this sample set the strongest agreement between assays was for IgG and total antibody (Pan or Bridging) responses on the same platform and with the same antigen (Figure 5D).

**Figure 5.**
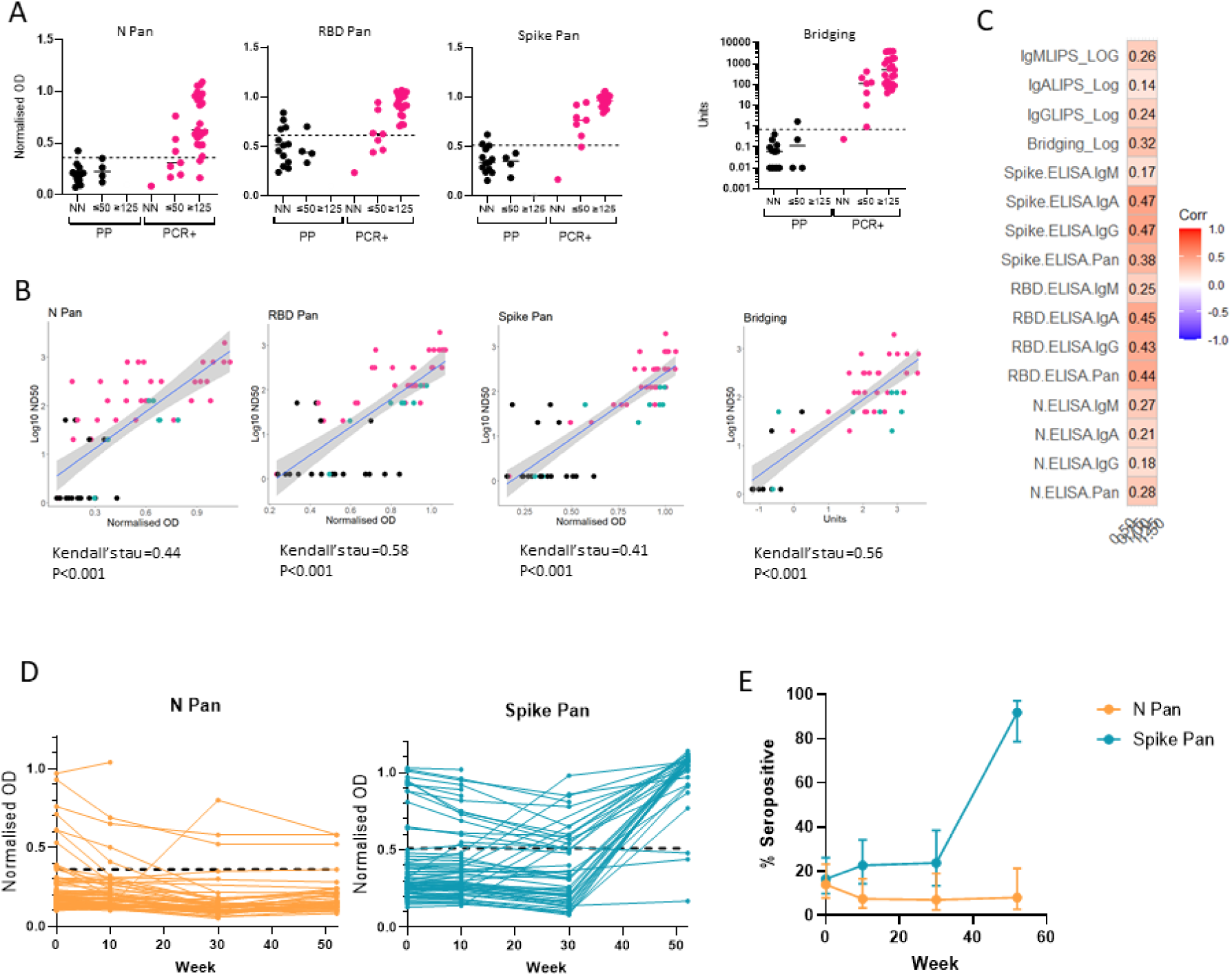
Relationship between binding antibody results and neutralisation titres and application of screening assay to longitudinal cohort. On a subset of the samples from the threshold and validation sets, we compared screening assay results to neutralising antibody titres were measured using a microneutralisation assay using SARS-CoV-2 and a pseudotype viral neutralisation assay using vesicular stomatitis virus (VSV) expressing Spike (A-C). (A) A microneutralisation assay was performed on 17 pre-pandemic serum samples and 31 from RT-PCR confirmed cases, and stratified the results into 3 groups: non neutralising (ND); ND50 of 20 or 50 (≤50); ND50 of 125 or above (≥125) and compared these groupings to the screening assay results: N Pan ELISA; RBD Pan ELISA; Spike Pan ELISA; Spike-RBD Bridging LIPS assay (where readouts are normalised OD or Units). (B) The relationship between results from each screening assay results and ND50 measured using the microneutralisation assay in n=59 samples displayed in scatterplots from a mixture of pre-pandemic (black), PCR-confirmed COVID-19 cases (pink) and exposed individuals or recent COVID-19 suspects (green). with a line showing the smoothed mean determined using a generalised linear model +/- 95% confidence intervals; and correlation performed using Kendall’s tau. (C) Correlogram showing the relationship between a novel pseudotype viral neutralisation assay (using mouse VSV expressing SARS-CoV-2 Spike and ACE-2 and TMPRSS-2) and all 16 ELISA and LIPS assays (total antibody and isotype specific) using n=38 COVID-19 samples from the validation set. (D) Field testing two screening assays (N Pan and Spike Pan ELISA) on longitudinal samples from a cohort of n=79 healthcare workers in Bristol in 2020 and 2021. (E) Observed seroprevalence/antibody positivity to N and Spike proteins using Pan ELISA assays in a cohort of n=79 healthcare workers. Total samples collected at each timepoint were as follows: week 0, n=79; week 10, n=66; week 30, n=42; week 52, n=37. 95% confidence intervals were calculated using the Clopper method.

### Reporting in international units

To ensure antibody levels measured using the in-house assays can be compared to those generated using other assays, we compared the WHO international standard to that of our in-house standard pool on a subset of the ELISAs (N Pan and IgG, Spike Pan and IgG). The WHO/NIBSC reference panel was used to compare antibody levels measured with our assays to the summary results using assays with similar antigen and antibody specificity from the inter-lab comparison study (19). Serial dilutions of both standards showed similar shaped curves when fitted to a 4PL regression and parallel lines (i.e. shared HillSlopes) for the two pools in all four assays (Figure 4E) thus allowing for conversion to international binding antibody units (BAU) values from the WHO standard to the in-house assays when using interpolation (Table S6); approximate BAU/ml values could also be assigned to the Norm OD thresholds. Furthermore, BAU/ml values identified on the in-house assay platforms were found to be comparable to the average values reported in the NIBSC inter-lab comparison (19), with the exception of samples with high levels of antibody, where the Spike Pan ELISA showed lower than expected levels, probably due to saturation of signal at this higher end of the assay signal range (Figure 4F).

### Exploring the relationship between binding antibody assay signals and neutralising activity

To identify any relationship between immunoassay results and the functional activity of antibodies in sera, assays were developed to measure virus neutralisation *in vitro* which were then deployed on a subset of our samples. A microneutralisation assay using SARS-CoV-2 virus was first used to measure virus neutralisation in samples from n=31 RT-PCR confirmed COVID-19 cases and n=17 pre-pandemic samples from the threshold set, with results expressed as half maximal dilutions (ND50) (Figure 5A,B). The pre-pandemic samples included some that had shown high signals in one or more of the immunoassays (i.e. false positives), to explore whether these false positives relate to functional cross-reactive responses. We observed low levels of virus neutralisation (ND50 <125) in only 4 of the 17 pre-pandemic samples, two of which displayed above-threshold immunoassay signals (one in the RBD Pan ELISA and another in the bridging assay). Amongst the samples from confirmed COVID-19 cases, samples with reasonable neutralising capacity (ND50 >125) had results in the high positive range for the Spike and RBD-specific assays (e.g. > 0.72 Normalised OD on the Spike Pan assay and >20 units on the Spike-RBD Bridging assay), but with the N Pan assay some strongly neutralising samples showing Normalised ODs around/below the optimal threshold for detection for an N-specific antibody response (Figure 5A). Agreement between the screening assay results and Log10 ND50 for neutralising samples from individuals with known or unclear status (as measured with the SARS-CoV-2 microneutralisation assay) was determined using Kendall’s rank correlation (Figure 5B). There was significant agreement in all cases, with the RBD Pan ELISA showing the highest tau coefficient (0.58). These results suggest agreement between the candidate screening assays results and functional capacity, but more so for the Spike-RBD specific assays that for the N Pan ELISA.

We also developed a pseudotype virus neutralisation assay using vesicular stomatitis virus (VSV) expressing SARS-CoV-2 Spike protein and VeroE6 cells expressing ACE2 and TMPRSS2 and deployed this on a larger set of samples from COVID-19 cases from across the sample sets representing a range of antibody responses using the full suite of assays (i.e. total antibody and isotype specific). There was good correlation between the neutralising titre values reported from the two assay platforms (Figure S5), but the pseudotype assays showed a wider dynamic range. Amongst the neutralising samples the Spike IgA and IgG ELISA assays showed the strongest correlation with half maximal neutralisation titre (Figure 5C).

### Field testing of screening assays

To field test two of the best-performing screening assays for determining changes in serological status in a population of interest during a time of vaccination rollout, the N and Spike Pan ELISAs were deployed on serum samples collected longitudinally from a cohort of n=79 hospital-based healthcare workers over a one-year period from April 2020 – May 2021 (Figure 5D). At baseline the rate of seropositivity with N Pan and Spike Pan was very similar at 13.92% and 16.46% respectively, but then the rate declined for N Pan to around 7% whilst remaining stable for Spike between weeks 10 and 30. By week 52 (end of study), we observed a divergence in the proportion of positive cases reported by the 2 assays, with Spike Pan showing up to 91.89% (78.70-97.20%) seropositive against Spike whilst there was negligible change in the proportion found to be seropositive to the nucleocapsid antigen (N) during the same period (8.11% (95%CI 280-21.30%), probably due to responses to vaccination among the majority of the cohort and low rates of new infections.

## Discussion

We optimised and evaluated a suite of in-house ELISA and LIPS assays for detecting and measuring antibody responses to SARS-CoV-2 by adapting available protocols (7, 14, 20). Whilst such assays have been used widely to explore the heterogeneity of immune responses to SARS-CoV-2 infection and vaccination (11, 12, 21), few have simultaneously and rigorously evaluated two different platforms with shared standards and quality controls using the same well characterised, large sample collections. This approach facilitated accurate threshold setting to determine serological status. The Spike Pan ELISA and Spike-RBD Bridging LIPS assays demonstrated superior performance in a blinded head-to-head comparison with a widely used commercial assay. All assays were optimised for low blood volume, low cost per sample and require relatively inexpensive laboratory equipment, paving the way for their use in a wide variety of settings and for a range of purposes, including population monitoring.

Initially the total antibody and isotype specific assays were compared for use as screening assays for recent SARS-CoV-2 infection in a subset of the threshold set. On both platforms, the assays measuring total antibody performed better than IgG, IgA or IgM specific assays in discriminating between samples from COVID-19 cases and pre-pandemic donors in the threshold sample set, and thus the four total antibody assays (N Pan, RBD Pan and Spike Pan ELISAs, and the Spike-RBD Bridging LIPS assay) were selected for validation as potential screening assays using a large, blinded validation set from pre-pandemic donors and confirmed/suspected COVID-19 cases. The superiority of the total antibody assays as sensitive tests for recent infection has been observed by others (22) and is unsurprising given the varying kinetics in antibody isotype responses over time, with a predominance of IgM and IgA isotypes in the acute stages followed by a dominant IgG response in most individuals after 21 days post infection (12). Importantly, our COVID-19 validation set included a wide spectrum of cases, including those with no or mild symptoms, who are likely to represent a large proportion of unknown community cases and clinically suspected hospitalised cases who may not have been tested for the virus at the optimal time. All total antibody assays achieved >97% specificity using at least one of the pre-defined thresholds. Sensitivity estimates for detection of recent COVID-19 cases varied but was highest for all assays when samples taken in late convalescence (>12 weeks post symptom onset) were used. The Spike Pan and Spike-RBD Bridging LIPS assays performed the best overall, providing up to 95% sensitivity for COVID-19 cases. Whilst not as sensitive as the two best performing screening assays, the N Pan and RBD Pan ELISAs provided a high level of specificity and could detect up to 82% and 73% of cases respectively. When sensitivity was compared directly to the commercial Roche Elecsys N assay, the Spike Pan ELISA and the Spike-RBD LIPS Bridging assays detected cases missed by the Roche assay and offered improved sensitivity despite using a 100-fold smaller volume of sample. A serology testing consortium (Oxford, UK) also reported a highly sensitive, specific and scalable anti-Spike IgG in-house ELISA which performed comparably to or better than commercial antibody assays in terms of diagnostic accuracy (23). Thus, in-house assays can offer high diagnostic accuracy and are suitable for population level surveillance.

We directly compared the ELISA and LIPS platforms by focusing on the RBD-specific antibody assays (as the antigen was common). On the ELISA platform, the RBD assays performed less well than the Spike assays. However, the detection of RBD-specific antibodies the novel Spike-RBD Bridging LIPS assay was better than the RBD Pan ELISA (sensitivities of 92.4% (95% CI 85.5-96.1) and 73.3 (95% CI 64.1-80.9), respectively, for infections confirmed by PCR >21 days previously). As has been reported elsewhere for COVID-19 bridging antibody assays, the Spike-RBD bridging LIPS assay results showed an upward trend in antibody levels over time since infection (24, 25) compared to a decline in levels observed with the corresponding RBD ELISA assays. It is likely that the bridging antigen format and the use of antigen competition for the IgG and IgA assays, select for higher affinity antibodies resulting in improved accuracy for discriminating between pre-pandemic and COVID-19 samples (25), but potentially also reflects affinity maturation due to somatic hypermutation in the weeks following priming. The bridging antigen format is used in other assays including the commercially available Roche Elecsys S and N assays but has not previously been applied to LIPS assays. LIPS has been found to offer improved diagnostic performance over other immunoassays including ELISA when deployed for detecting specific antibodies involved in identifying cases of autoimmune or infectious diseases (26). When measuring antibody levels in one sample dilution, the LIPS platform also provides a broader dynamic range. In contrast to ELISA, the isotype-specific LIPS assays should not be affected by within-well competition from other antibody isotypes, which may also explain the differences observed in this comparison. In conclusion, we found LIPS assays to be highly sensitive for the measurement of SARS-CoV-2 RBD specific antibodies, which may offer unique information on the affinity of antibody binding when compared to other platforms. Further investigation into why antibody kinetics are profiled differently for same samples measured using assays from different platforms will inform the optimal use of each platform for a given purpose relating to the measurement of specific antibody (e.g. as a proxy for a functional responses versus to accurately quantify antibody decay rates).

ELISA and LIPS assays measure levels of binding antibody to specific antigens, but these measurements do not necessarily correlate accurately with antibody-mediated protection. Neutralisation assays are widely deployed to monitor functional antibody responses to SARS-CoV-2 and nAb levels have been shown to correlate with protection against re-infection and/ after vaccination (27). However, in this study and others, binding antibody levels (particularly IgG/total antibody to the RBD and/or Spike antigens) correlated strongly with nAb levels when measured using two different neutralisation assays (28), and have been shown to correlate with protection (4). Thus, to some extent, binding antibody assays specific to RBD and Spike can be used as surrogates for functional, protective antibody responses. Further work may lead to the identification of binding antibody thresholds of our candidate assays indicative of protection and/or sufficient levels of nAbs. We identified some false positive ELISA/LIPS samples in the pre-pandemic sample sets, and 4 samples with low level neutralising capacity, which could reflect the presence of cross-reactive antibodies in these samples perhaps resulting from other CoV infections. Interestingly, there was little agreement across the different assays in terms of which samples were identified as positive amongst pre-pandemic samples, even among those that were tested by neutralisation -i.e. not all with neutralising capacity demonstrated binding to RBD/Spike. This suggests that false positive pre-pandemic samples were not the result of cross-reactive antibody responses. Unlike Ng et al, who reported that antibodies from pre-pandemic samples had binding capacity to the S2 portion of Spike (17), we did not observe binding to Spike in the 4 pre-pandemic samples with low neutralising capacity.

We demonstrate the feasibility of assigning international BAU to samples measured with our assays, and report similar levels of antibody to other laboratories who contributed to the development of international reference standards (19, 29), allowing comparison of results from our assays with others. We deployed two of the screening assays (Spike and N Pan ELISA) on samples from a longitudinal cohort of healthcare workers in Bristol, UK (LOGIC). These data suggest comparable sensitivity for recent infection prior to vaccination rollout in the UK but, as expected, an increase in anti-Spike specific responses was observed after vaccination was introduced and the seropositivity rate was very high (>95%) by the end of the 12 month follow up period (31st May 2021), causing a divergence in the relative seropositive rates within the cohort. As such, using a combination of our assays to measure antibody positivity rates in order to estimate recent infection and/or vaccination exposures is feasible. In countries/areas with high vaccination coverage, the nucleocapsid specific assays are therefore increasingly of relevance for seroprevalence studies, whilst Spike/RBD assays (as well as neutralising antibody assays) are less able to differentiate vaccination-specific from infection-specific immunity, but are probably better correlates of immunity. However, not all vaccines are Spike-only (e.g., the Valneva vaccine contains the whole virion), and the relative rate of seroconversion to nucleocapsid after prior exposure to Spike via vaccination (compared to naïve individuals) has not been ascertained. Thus, there are important considerations to bear in mind when using nucleocapsid assays to determine recent infection amongst vaccinated individuals.

Strengths of this study include rigorous development of high performance, low blood volume, cost-effective tests which can be easily deployed in a variety of settings, but our approach also has several limitations. Firstly, whilst samples from pre-pandemic children were included, samples from children with COVID-19 were not available to us and as such, assay performance for detecting recent paediatric infections cannot be reported. However, since widespread vaccination of children is not currently common in many countries while asymptomatic/mild paediatric infections are, antibody assays offer a useful tool for monitoring infection in this age group. The antigens used in the in-house assays were generated using the genetic sequence from the parent Wuhan strain of SARS-CoV-2 first described in 2020 (7) from which several new variants of concern (VOC) have evolved and have caused significant waves of infection globally. Some of these variants, especially Omicron, include multiple mutations in these target antigens and as such, may lead to antibody responses with differential binding to the target antigens. Indeed, antibodies responses raised to antigens from one SARS-CoV-2 variant genetic sequence lead to differential ability to neutralise VOC strains. However, whilst others have shown reduced binding to antigens from sequences of VOCs, rates of seropositivity when using different antigens, and/or from people who were infected with non-Wuhan variants, appear to be relatively unchanged (30, 31).It will be important to monitor changes in antibody assay performance for detecting recent infection as new variants emerge and become dominant.

In summary, we present a blueprint for the development and evaluation of low volume antibody assays for screening for seropositive individuals and/or profiling of serum isotype-specific antibody responses after SARS-CoV-2 infection and/or vaccination. With the use of appropriate controls, these assays offer a low cost and reliable alternative to commercial assays and can be used with simple laboratory equipment, potentially allowing for infection/immunity monitoring in hard-to-reach communities (via field or at-home sampling) and in low-income settings where other testing approaches are not feasible. Evaluation of one or more of these assays in areas where epidemiological information on COVID-19 is sparse, could be of great value.

## Methods

### Serum and Plasma Sample collection

Blood was collected into SST vacutainers (BD Biosciences) and serum was separated by centrifugation at 1300 x *g* for 10 minutes, before being aliquoted into cryovials and stored at -70°C.

### Heat Inactivation

Samples from suspected and proven PCR confirmed COVID-19 cases were heat treated to inactivate live virus, at 56°C for 30 minutes according to local and national health and safety guidance.

### Antigen Production

#### SARS-CoV-2 trimeric Spike protein, and Receptor Binding Domain (RBD)

SARS-CoV-2 Spike ectodomain and RBD were expressed in Hi5 insect cells as previously described (7) . The Spike ectodomain construct was comprised of amino acids 1 to 1213 fused with a thrombin cleavage site followed by a T4-foldon trimerization domain and a hexahistidine affinity purification tag at the C-terminus. The polybasic furin cleavage site was mutated (RRAR to A) (7). The construct for RBD was comprised of amino acids 1-14 (secretion signal) and then 319-541 of the Spike ectodomain and was followed by a hexahistidine affinity purification tag at the C-terminus (7, 32). Both Spike protein and RBD were purified using a previously described purification protocol (33). Briefly, supernatant media containing expressed proteins were harvested from transfected cells 3 days post-transfection and were incubated with HisPur Ni-NTA Superflow Agarose (Thermo Fisher Scientific) for 1h at 4°C. Resin bound with SARS-CoV-2 Spike or RBD protein was separated and extensively washed with wash buffer (65 mM NaH_2_PO_4_, 300 mM NaCl, 20 mM imidazole, pH 7.5) and finally protein was eluted with buffer (65 mM NaH_2_PO_4_, 300mM NaCl, 235mM imidazole, pH 7.5). Eluted protein was then concentrated, and buffer exchanged to phosphate-buffered saline (PBS) pH 7.5 using 10 kDa (for RBD) and 50 kDa (for Spike) MWCO Amicon centrifugal filter units (EMD Millipore). Concentrated proteins were then aliquoted, flash frozen in liquid nitrogen and stored at -80°C until further use.

#### Nucleoprotein (ELISA)

The *E. coli* codon optimised nucleotide sequence of full-length nucleoprotein from SARS-CoV-2 was synthesized by GenScript. The sequence was synthesized with an *NdeI* restriction site at the 5’ end and the *BamHI* site at the 3’ end and cloned into pET28a expression vector. All proteins were expressed with C-terminal His_6_-tags to facilitate subsequent purification. The recombinant plasmids (pET28a-NP-FL) were transformed into *E. coli* strain BL21 (DE3). Protein expression was induced by the addition of 1 mM IPTG and then incubated overnight at 20°C. Cells were pelleted by centrifugation and resuspended in 20 mM Tris pH 8, 500 mM NaCl, 10 mM imidazole, 1 mM NaF and 1 mM PMSF. Cells were lysed by passage through a French Press cell (Spectronic Instruments) and the resulting lysates were centrifuged at 39,000g at 4°C for 30 min. The supernatant was applied to a HisTrap HP nickel affinity column (GE Healthcare) and washed with a series of gradient wash buffers (20 mM Tris pH 8, 500 mM NaCl, 10, 20 and 40 mM Imidazole). The protein was eluted in 20 mM Tris pH 8, 500 mM NaCl and 500 mM imidazole and further purified by size exclusion chromatography using a HiLoad 16/600 Superdex 200 ® pg column (GE Healthcare) equilibrated and eluted in 20 mM Tris pH 8 and 500 mM NaCl. Peak fractions were pooled and concentrated in a 10 kDa MWCO Vivaspin ultrafiltration unit. Protein concentration was determined using the Bradford assay. Typical yields of N proteins after Ni-NTA and size exclusion chromatography was approximately 9 mg/L. Purified proteins were analysed by SDS-PAGE and by Western-blots assays using an anti-His tag antibody (Sigma).

#### Luciferase tagged RBD for LIPs

RBD antigen tagged with a luciferase reporter (nanoluciferase, Nluc) was generated and provided by Dr Vito Lampasona (Milan) for a Luciferase Immunoprecipitation System (LIPS) assay (14, 33).

### Production of in-house standard and controls

To allow for assay standardisation and to monitor quality control of results over time, including intra- and inter-plate variability, control material was generated from large volume serum samples with known recent infection and well-characterised antibody status. For the pooled standard, (which was used for all ELISA assays, the RBD LIPS Bridging and IgG assays) sera from 3 mildly affected PCR-confirmed cases were combined and aliquoted.

### ELISA

The ELISA protocol is based on the RBD screening and Spike confirmatory ELISA assays described in (7, 20) with some modifications. MaxiSorp high-binding ELISA plates (NUNC) were coated with Spike (10 µg/ml), RBD (20 µg/ml) or N protein (20 µg/ml) in PBS and incubated overnight at 4°C (except for IgM ELISAs, where all antigens were coated at 2 µg/ml). Subsequent steps were all performed at room temperature (RT). Unbound antigen was removed with 3x washes in PBS with 0.1% v/v Tween-20 and plates were blocked for 1 hour with 3% BSA/PBS or 3% milk (Sigma) for IgM assays. Serum samples were diluted in either 1% BSA/PBS or 1% milk (for IgM only) and incubated on the plate (100 µl per well) for 2 hours. After washing, HRP-conjugated anti-human Pan-Immunoglobulin (Pan) (Sigma), IgG (Southern Biotech), IgA (Sigma) or IgM (Sigma) secondary antibody, in the same dilution buffer as the samples, was added (50 µl per well) and incubated for 1 hour. Plates were washed and dried before development with the HRP substrate OPD (SigmaFast; 100 µl per well). The reaction was terminated after 30 minutes by the addition of 3 M HCl (50 µl per well). Optical density was measured at 492 nm and 620 nm on a BMG FLUOstar OMEGA MicroPlate Reader with MARS Data Analysis software. The 620 nm reference wavelength measurements were subtracted from the 492 nm wavelength measurements for each well to give background corrected values. Averaged blank values from wells containing no serum were then subtracted from all experimental values for each plate.

Three read out approaches were explored for test samples: OD values normalised to the internal positive control, an area under the curve (AUC) generated from a 4-point dilution series (where available), and an interpolated value (arbitrary units).

### LIPS

Non-competitive: The Nluc-RBD antigen (5) was diluted in 20mM Tris, 150mM NaCl, pH 7.4 with 0.5% v/v Tween-20 (TBST) and 0.05% casein to 4x10^6^+/-5% per 25µl. All steps were carried out at RT, unless specified. Reagents were stored at 4°C, TBST and diluted antigen were used directly from 4°C, Nano-Glo® substrate was equilibrated to RT before use. Serum samples (1µl, 2 replicates) were pipetted into a 96-well plate and incubated with 25µl diluted antigen for 2 hours in a dark area. Immunocomplexes were precipitated using 2.5µl glycine-blocked Protein A Sepharose 4 fast flow (GB-PAS) (GE Healthcare Life Sciences, Chicago, IL, USA) and 2.5µl ethanolamine-blocked Protein G Sepharose (EB-PGS) (GE Healthcare Life Sciences) (washed 4 times in TBST) for 1hr with shaking (∼700rpm). Precipitates were washed 5 times with TBST and then transferred to a 96-well Optiplate^TM^ (Perkin-Elmer, Waltham, MA, USA) and excess buffer removed by aspiration. Nano-Glo® substrate (40µl, Promega) was injected into each well immediately before counting in a Hidex Sense Beta (Hidex, Turku, Finland).

Competitive: To overcome cross reactive responses from non-COVID-19 samples, SARS-CoV-2 RBD protein (as used in ELISA) was used to outcompete the Nluc-RBD label. A range of concentrations of RBD were tested with 8x10^-8^ mol/L showing good affinity for RBD specific IgG. Sera (1µl, 4 replicates) were pipetted into a 96 well plate. The Nluc-RBD antigen was diluted in TBST with 0.05% casein to 4x10^6^+/-5% LU per 25µl with or without unlabelled RBD added at a final concentration of 8x10^-8^ mol/L. Two replicates of each samples were incubated with Nluc-RBD and two replicates with competition of Nluc-RBD binding with unlabelled RBD. Immunocomplexes were precipitated and measured as outlined above. Where antibodies were of higher affinity unlabelled RBD outcompeted binding of Nluc-RBD, lowering the LU measured. A delta LU was calculated (mean LU of uncompleted wells - mean LU of competed wells) and then interpolated from LU by the standard curve, creating LIPS units corrected for non-specific binding.

IgA and IgM measurement: IgA was measured in 1µl serum per replicate, with competitive displacement (as for IgG) and immunoprecipitated using 3.75µl per well IgA agarose (Sigma), in place of GB-PAS/EB-PGS. IgM was measured in 2µl serum per replicate, without competitive displacement using 5µl per well IgM agarose (Sigma).

#### Spike-RBD Bridging LIPS

To develop a novel LIPS bridging assay format for high throughput requirements, Spike antigen diluted to 100ng/40µl in PBS was pipetted into every well of a 96-well high-binding OptiPlate™ (Perkin-Elmer, Waltham, MA, USA) and incubated for 18hrs at 4°C. The plate was washed 4 times with TBST and blocked with 1% Casein in PBS (Thermo Scientific, Waltham, MA, USA). The plate was left to air-dry for 2-3hrs before being stored with a sachet of desiccant in a sealed plastic bag at 4°C and used within three weeks.

The Nluc-RBD antigen was diluted in TBST to 10x10^6^+/-5% LU per 25µl. Sera (1.5µl, 2 replicates) were pipetted into a 96-well plate and incubated with 37.5µl diluted labelled antigen for 2hrs. Of this mixture, 26µl was transferred into the coated OptiPlate and incubated shaking (∼700rpm) for 1.5hrs. The plate was washed 8 times with TBST, excess buffer was removed by aspiration, then 40µl of a 1:1 dilution of Nano-Glo® substrate (Promega) and 20mM Tris 150mM NaCl pH 7.4 with 0.15% v/v Tween-20 was injected into each well before counting in a Hidex Sense Beta Luminometer (Turku, Finland). Units were interpolated from LU through a standard curve.

#### Roche SARS-CoV-2 anti-nucleocapsid antibody assay

Serum samples from PCR-confirmed cases were analysed using the commercial Elecsys^®^ Anti-SARS-CoV-2 (Roche) in the Department of Microbiology, Infection Sciences, Southmead Hospital, North Bristol NHS Trust, Southmead Road, BS10 5NB, UK following manufacturer’s instructions. Elecsys® Anti-SARS-CoV-2 is an immunoassay for the in vitro detection of total antibodies (including IgG) to SARS-CoV-2 in human serum and plasma. The assay uses a recombinant protein representing the nucleocapsid (N) antigen in a double-antigen sandwich assay format, which favours detection of high affinity antibodies against SARS-CoV-2.

### Microneutralisation assay

VeroE6 cells (ATCC) were cultured in Dulbecco’s Modified Eagle’s medium containing GlutaMAX (Thermo Fisher Scientific) supplemented with 10% fetal calf serum (FCS) (Thermo Fisher Scientific) and 0.1 mM non-essential amino acids (NEAA) (Sigma Aldrich) at 37°C in 5% CO_2_. For immunofluorescence analysis, cells were seeded the day prior to infection in appropriate media in µClear 96-well Microplates (Greiner Bio-one). Neutralising capacity of human serum samples was quantified using a microneutralisation assay as previously described (33). Briefly, heat-inactivated serum was serially diluted 2.5-fold from 1:20 for 8 dilutions and incubated with live virus (SARS-CoV-2/human/Liverpool/REMRQ0001/2020) for 60 mins at 37°C. Following incubation, the mixtures of virus and diluted sera were added to VeroE6 cells and incubated for 18 hours before being, fixed and stained with antibodies against the SARS-CoV-2 N protein (1:2000 dilution; 200-401-A50, Rockland) followed by an Alexa Fluor-conjugated secondary antibody (ThermoFisher) and DAPI (Sigma Aldrich). Images were acquired on the ImageXpress Pico Automated Cell Imaging System (Molecular Devices) using a 10X objective and infected cells detected and quantified using Cell ReporterXpress software (Molecular Devices). The percentage of infected cells was calculated relative to control wells which contained virus only, without serum.

### Pseudovirus neutralisation assay

Luciferase-expressing vesicular stomatitis virus (VSV*ΔG-FLuc particles) were a gift from Yohei Yamauchi. VSV-G-harbouring BHK21 cells were infected with VSV*ΔG-FLuc particles to generate complemented VSV*G-FLuc particles as previously described (34). To generate Spike-harbouring pseudovirus (VSV-S-FLuc), 293T cells were seeded and transiently transfected with a plasmid corresponding to the original Wuhan strain Spike protein (pCAGGS-S2-spike) using Turbofect transfection reagent (ThermoFisher R0532) for 16 hours following the manufacturer’s instructions. Transfected 293T cells were then infected with VSV*G-FLuc particles for 2 hours, washed with PBS, then incubated with fresh DMEM, supplemented with 10% FBS and 1:2000 (v/v) I1 (anti-VSV-G) antibody (absolute antibody Ab01401-10.3). Optimal pseudotype cell entry was achieved using VeroE6 cells stably expressing the human angiotensin-converting enzyme 2 (ACE2) receptor and the cell surface protease TMPRSS2 (Vero ACE2 TMPRSS2 (VAT) cells, which were a kind gift from Dr Suzannah Rihn, MRC-University of Glasgow Centre for Virus Research (35).

For pseudovirus neutralisation assays, 10,000 VAT cells were seeded per well in opaque, white 96-well plates. The following day, serum samples were titrated 2.5-fold, 9 times across 96-well plates from a starting dilution of 1:40. Pseudovirus corresponding to 10,000 RLU was immediately added to each well, mixed and incubated for 1 hour. After aspirating cell media from VAT cells, pseudovirus/serum and control mixtures were added to corresponding wells on VAT cells and incubated at 37°C 5% CO_2_ overnight. Luminescence measurements were taken 16 hours after infection, using the ONE-Glo Luciferase Assay System (Promega). Luminescence was measured using a Tecan Infinite 200 plate reader at room temperature.

### Blinding of validation set

The validation set of samples (n=807) were split into multiple aliquots (n=5) for randomisation and blinding by assigning a new barcode ID for each aliquot. The 5 sets of samples each had a unique order and the LIPS and ELISA assays were performed one separate sets, such that each lab remained blind to the results of the other. Screening ELISAs were performed on the same samples at the same time. Unblinding took place after all assay were performed and the data had been finalised.

### Statistical Analysis

Data analyses were performed using either R software with R Studio, and GraphPad Prism (version 9) as detailed below.

#### Standardisation to serum pool standards

In both ELISA and LIPS assays, each plate or set of 2 plates included a dilution series of the in-house serum pool, to standardise sample values and control samples across plates. For the ELISA assay, sample values are reported as normalised ODs, where average OD values are divided by the top standard value in the plate. However, sample readouts using other methods including interpolated unit values (from a 4-parameter logistic regression model fit (on Prism or within BMG software) to the 7-point standard pool dilution series) and AUC from sample dilution series were used in the development stage. For LIPS assays, the raw luminescence unit values for the standard pool dilution series across two plates were fitted to a logarithmic curve to allow for interpolation of in-house unit values to average raw light unit values for samples.

The WHO International Standard (First WHO International Standard for anti-SARS-CoV-2 immunoglublin (human) (NIBSC code 20/136) and the WHO Reference Panel (First WHO Reference Panel for anti-SARS-CoV-2 immunoglobulin) (NIBSC Code 20/268) were both purchased from the National Institute for Biologic Standards and Control (NIBSC), Potters Bar, Hertforshire, UK in 2021.

#### Diagnostic accuracy estimates and setting of thresholds

Performance of candidate tests for discriminating between pre-pandemic and COVID-19 samples was performed using Receiver Operator Characteristic (ROC) Curve analysis. From such analysis of threshold set samples, thresholds meeting the following criteria were identified and selected for evaluation in the blind evaluation: 99^th^ centile of pre-pandemic levels (to achieve 99% specificity); 98^th^ centile of pre-pandemic samples and the point at which the highest Youden’s index is achieved (Jmax). In selected samples sets based on recommended categorisation of COVID-19 cases, sensitivity and specificity at target thresholds were determined using ROC curve analysis and, where appropriate, estimates on sensitivity and specificity were reported at a pre-specified threshold; 95% confidence intervals around these estimates were calculated using the Clopper method without correction for multiple comparisons.

#### Assessing for parallel lines

To assess for parallelism between the in house and international serum standards, a full standard curve (in duplicate) of the in-house pooled standard was ran on the same plate as the WHO international standard (NIBSC code 20/136 as above), at the appropriate dilution range for each assay. This was repeated up to 3 times to ensure robust comparisons and to account for inter-plate variability. Duplicate ODs from each dilution in each series were averaged, and plate replicates were combined as repeats for each standard and fitted to a non-linear regression model (usually 4-parameter logistic) curve. The combined dilution series for each standard (where average ODs from each plate were classed as replicate values) were used to assess for goodness of fit of two nested models: one where the HillSlope of the models is shared, and one where they differ. The resulting F statistic and P value determine whether the Model where HillSlope is shared (i.e. the lines are parallel for the midpoint).

#### Assigning BAU values to in house standards

For those assays where the international standard and in-house standard were found to be parallel, replicated run values were used to ascertain the IC50 values for each standard, followed by the ratio between the control samples.

#### STARD Checklist

This study conforms to the STARD checklist for publishable diagnostic accuracy studies (36), as outlined in Table S7.

### Study approval

This work utilises various collections of samples collected both before and during the COVID-19 pandemic via different protocols. Details are indicated in 3 sections below.

#### COVID-19 cases

PCR confirmed and clinically suspected COVID-19 cases were recruited via two independent routes in Bristol, United Kingdom between April and November 2020 (prior to roll out of vaccinations). PCR confirmed and clinically suspected severe COVID-19 cases admitted to hospital were recruited into the DISCOVER study at North Bristol NHS Trust for which HRA Approval was granted by the South Yorkshire Research Ethics Committee (20/YH/0121). Clinical and demographic features were recorded during the in-patient stay and subsequently at out-patient follow-up clinics. Respiratory samples were submitted for SARS-CoV-2 PCR testing on admission or first presentation; further samples were tested where initial samples tested negative for SARS-CoV-2 RNA. Serum or plasma samples were collected at various time points during in-patient and out-patient follow-up.

In addition, healthcare workers (HCWs) at University Hospitals Bristol, North Bristol NHS Trust and Weston NHS Foundation Trust who had a previous positive nasopharyngeal swab for SARS-CoV-2 by PCR were invited to donate blood samples under the Bristol Biobank (NHS Research Ethics Committee Ref 20/WA/0273). A clinical data form was used to collect details of symptoms, tests and other information relating to the donor and their COVID-19 status at the point of sampling. A subset of donors was invited back for a repeat donation at >12 weeks post symptom onset.

#### Pre-pandemic samples

Samples collected prior to December 2019 were sourced from various collections held in Bristol as detailed in the Table 3 below. All samples were used in accordance with the Human Tissue Act (2004) with appropriate consent and ethical approvals in place.

**Table 3.**
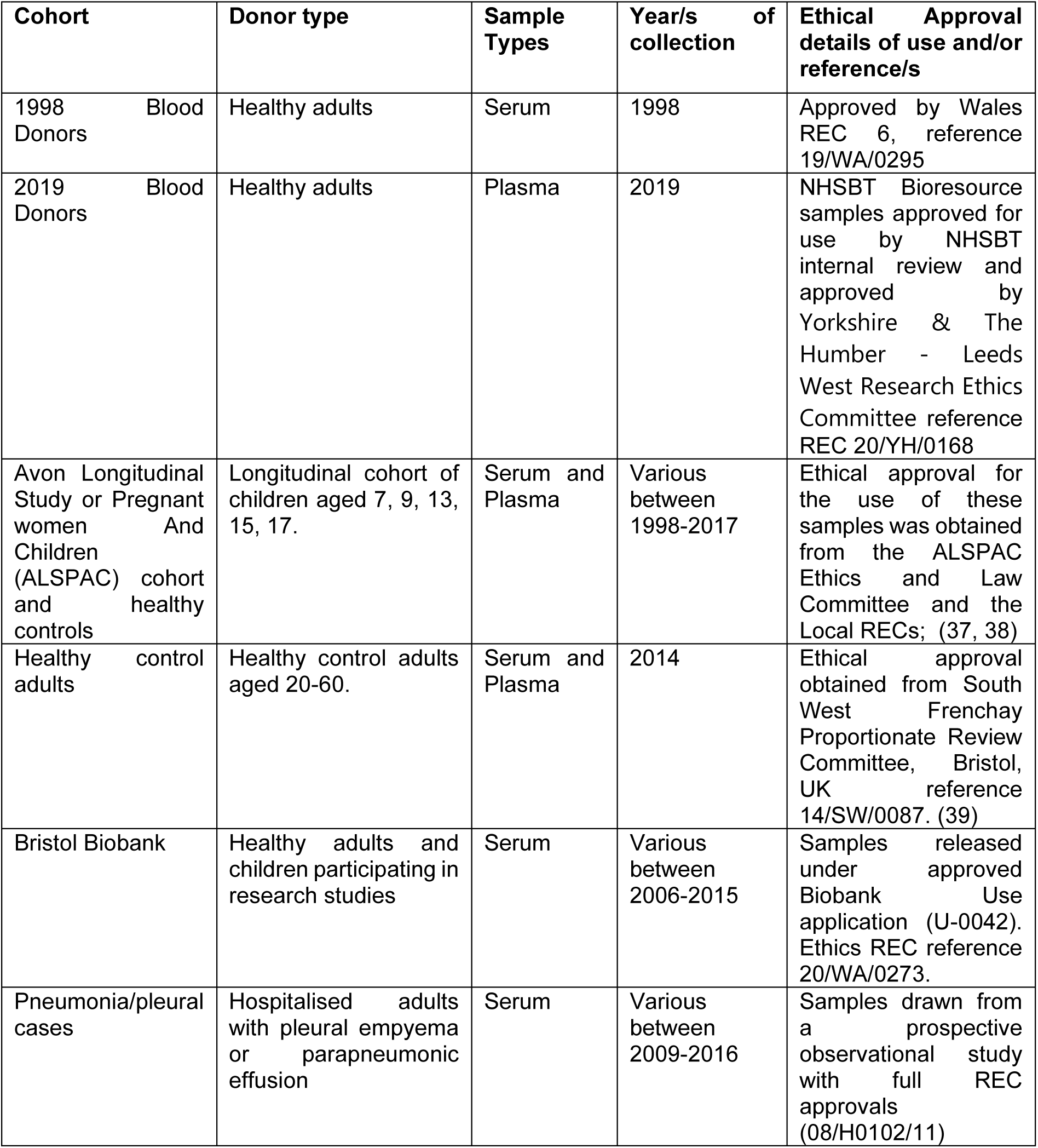
Details of pre-pandemic samples collated from various collections including research ethics committee (REC) references, sample types and years of collection.

#### Samples from a longitudinal cohort of hospital-based healthcare workers collected during the pandemic

Patient-facing clinical staff members working in the Children’s Emergency Department (CED) of Bristol Royal Hospital for Children, including doctors, nurses and healthcare assistants were invited to take part in the LOGIC (<u>LO</u>n<u>GI</u>tudinal Study of <u>C</u>OVID-19: Symptoms, Virology & Immunity) study in April 2020; REC reference number 20/YH/0148. In this study, participants donated blood samples over a 12-month period.

### Author contributions

AH, AEL, HEB, ACT, AF, DNW, MB, AJKW & KMG conceived/designed the study. AG, JM, RB, CP, FH & DA recruited recently infected SARS-CoV-2 participants/donors and healthcare workers. EO, MG, LK, AH, HEB, LC, JO, BMA, LW, LR, SR, NT, FH, DA, AT, KC & KMG sourced, provided and/or processed samples. KG, NB, AT, IB produced antigens. HEB, AH, EO, BH, JS & UO performed the ELISAs. MR, YBK, IK, GM, OB & KC performed the LIPS labwork. MS, LP & VL provided LIPS reagents. JM & PM performed the Roche Elecsys nucleocapsid bridging assay. OF, MKW, LW & ADD developed and performed the pseudoneutralisation and neutralisation assays. AH, AEL, HEB, CP, ACT & KLS performed the data analysis. AH, AEL, HEB, KLS & KNG wrote the manuscript. All authors read and approved the final manuscript.

## Data Availability

All data produced in the present study are available upon reasonable request to the authors

## Acknowledgements

We dedicate this manuscript to our colleague and friend, Alistair Williams, who sadly passed away in September 2020 after a brave battle with cancer. Alistair was an expert in autoantibody assay development and used this expertise to guide us through incorporation of optimal standardization and quality control approaches.

We are extremely grateful to all the donors (COVID-19 cases and pre-pandemic donors) who provided serum and plasma samples for scientific research; this project highlights the value of Biobank/sample collections for scientific research and development. For those samples from the ALSPAC study, we also thank the midwives for their help in recruiting the study participants, and the whole ALSPAC team, which includes interviewers, computer and laboratory technicians, clerical workers, research scientists, volunteers, managers, receptionists, and nurses.

We thank Hayley Jones for guidance on the statistics and analytical approach. We thank Jill King for support in recruitment of Biobank donors, and Helen Thompson for support in coordinating sample donations via the Biobank, and for wider administrative support. We thank Leigh Johnson and Doug Owen for supporting sample collection for the LOGIC study. We are grateful for the Bristol UNCOVER community for advice, guidance and support over the past 2 years.

## Data availability statement

The supporting data relating to this manuscript will be made available via an online repository at the point of the article being accepted in a journal after peer review. Until then, all supporting data files are available upon request from the corresponding author

