## Supplementary material for "Development and evaluation of low-volume tests to detect and characterise antibodies to SARS-CoV-2"

**Table S1. Full threshold setting cohort.** Key clinical and demographic features of serum/plasma donors in the Threshold Setting Set (stratified by case status i.e. 'Pre-pandemic' and 'COVID-19').

| | **Pre-pandemic** | **COVID-19 cases** |
|---|---|---|
| Total | 399 | 47 |
| Sample Type, n (%) | | |
| Serum | 325 (80.9) | 38 (80.6) |
| Plasma | 74 (19.1) | 9 (19.1) |
| Baseline | 399 (100.0) | 44 (93.6) |
| Repeat | 0 | 3 (6.4) |
| Age group: | | |
| Children 4-9 years, n (%) | 61 (15.4) | 0 (0.0) |
| Teenagers 13-17 years, n (%) | 30 (7.44) | 0 (0.0) |
| Adult >18 years, n (%)[&] | 308 (77.2) | 47 (100) |
| Gender: | | |
| Male, n (%) | 135 (33.5) | 21 (44.7) |
| Female, n (%) | 139 (40.0) | 25 (53.2) |
| Unknown, n (%) | 129 (9.7) | 1 (2.1) |
| Cohort and collection period: | | |
| Child research study (2014), n (%) | 1 | 0 |
| ALSPAC (1998-2010), n (%) | 100 | 0 |
| ALSPAC controls (2014), n (%) | 48 | 0 |
| Blood donors (1998 and 2019), n (%) | 253 | 0 |
| Hospitalised COVID-19 cases (2020), n (%) | 0 | 16 |
| Community COVID-19 cases (2020), n (%) | 0 | 31 |

& - Most of those with unknown exact age were part of the NBS Plasma cohort, who must by nature be adults as that is a requirement for blood donation. N=1 was a lab donor control from ALSPAC and must also be an adult.

**Table S2. Selection of thresholds for screening assays and corresponding performance in threshold set.**

| **Assay name** | | **N Pan (ELISA)** | **RBD Pan (ELISA)** | **Spike Pan (ELISA)** | **Spike-RBD Bridging LIPS** |
|---|---|---|---|---|---|
| PCR positive/convalescent (n) | | 47 | 47 | 47 | 46 |
| Pre-pandemic | | 399 | 399 | 399 | 401 |
| Read-out | | Normalised OD | | | Units |
| AUC (95% CI) | | 0.947 (0.903-0.99) | 0.945 (0.898-0.992) | 0.947 (0.889-1.00) | 0.997 (0.993 to 1.001) |
| Threshold 1<br><br>99th percentile | Threshold (units) | 0.40 | 0.70 | **0.51** | 0.45 |
| | Sensitivity (95%CI) | 74.47% (60.49% to 84.75%) | 76.6% (62.78% to 86.40%) | 91.49% (80.07% to 96.64%) | 95.65 (85.16% to 99.47%) |
| | Specificity (95% CI) | 99% (97.45% to 99.61%) | 99% (97.45% to 99.61%) | 99% (97.45% to 99.61%) | 98.75 (97.11% to 99.59%) |
| Threshold 2<br><br>98th percentile | Threshold (units) | **0.36** | 0.67 | 0.49 | 0.35 |
| | Sensitivity (95%CI) | 78.72% (65.10% to 88.01%) | 78.72 (65.10% to 88.01%) | 91.49% (80.07% to 96.64%) | 95.65 (85.16% to 99.47%) |
| | Specificity (95% CI) | 97.99 (96.09% to 98.98%) | 97.99 (96.09% to 98.98%) | 98.75% (97.10% to 99.46%) | 97.76 (95.78% to 98.97%) |
| Threshold 3<br><br>Highest Youden's Index | Threshold (units) | 0.31 | **0.61** | 0.485 | **0.8** |
| | Sensitivity (95%CI) | 82.98% (69.86% to 91.11%) | 82.98% (69.86% to 91.11%) | 93.62% (82.84% to 97.81%) | 95.65 (85.16% to 99.47%) |
| | Specificity (95% CI) | 96.99% (94.82% to 98.27%) | 97.24% (95.13% to 98.45%) | 97.99 (96.09% to 98.98%) | 99.75 (98.62% to 99.99%) |

**Table S3. Inter- and intra-assay variation of the screening assays deployed on the full validation set, using data from standards and controls.**

| | | Inter-assay coefficient of variation | | | Intra-assay coefficient of variation | | | |
|---|---|---|---|---|---|---|---|---|
| **Assay** | **Number of plates** | **Low Positive** | **High Positive** | **Negative** | **Top standard** | **Low Positive** | **High Positive** | **Negative** |
| N Pan ELISA | 10 / 20 * | 4.09 $ | 13.19 | 14.69 | 2.12 | 2.96 $ | 2.71 | 3.68 |
| RBD Pan ELISA | 10 / 20 * | 16.01 | 1.91 | 18.53 | 2.45 | 4.16 | 2.58 | 5.43 |
| Spike Pan ELISA | 10 / 20 * | 11.35 | 1.8 | 9.82 | 1.74 | 2.40 | 2.39 | 3.14 |
| Spike-RBD Bridging LIPS | 15,31,28,26/31# | 23.39 | 10.85 | 180.41 | 8.98 | 9.41 | 5.84 | 25.00 |

* Top standard and negative control were run on every ELISA plate, low and high positive controls we run once per pair of plates

### for Top standard, Low, High, Neg (respectively). QCs run once per plate and standards once per pair of plates but some QCs were rejected and samples affected were repeated.

$ low positive data from one ELISA plate removed from these calculations as results were outside of the accepted range; other QCs and standards on these plates were within the defined ranges and therefore the data on the plates was accepted.

**Table S4. Screening assay sensitivities for detection of n=139 Samples from PCR-confirmed individuals collected >21 days post symptom onset/PCR test (a sensitivity analysis).** Within the validation set, 149/222 of the COVID-19 case samples were from PCR- confirmed cases and at least 21 days post symptom onset or positive PCR test. After removal of the 'suspected' cases and samples taken between 0-21 days post symptom onset, n=13 repeat samples (first samples for each donor was included) were removed – with ??? samples remaining. This cohort was used to calculated the sensitivities of candidate screening assays for PCR-confirmed individuals >21 days symptom onset (i.e in those where an antibody response is expected).

| | **Sensitivity for PCR confirmed COVID-19 >21 days p.s.o at defined thresholds (95% CI)** | | | **Optimal Overall Threshold (method)** |
|---|---|---|---|---|
| Threshold method* | 1 | 2 | 3 | |
| N Pan ELISA | 70.5 (61.1 - 78.4) | 77.1 (68.1 - 84.2) | 81.9 (73.3 - 88.2) | (2) |
| RBD Pan ELISA | 63.8 (54.2 - 72.4) | 65.7 (56.1 - 74.2) | 73.3 (64.1 - 80.9) | (3) |
| Spike Pan ELISA | 93.83 (90.12-96.93) | 94.44 (87.8 - 97.4) | | (1) |
| Spike-RBD Bridging LIPS | 92.4 (85.5 - 96.1) | | | (3) |

*Same as reported in Table 2

**Table S5. Sensitivity of screening assays compared to Roche Elecsys N in different categories of COVID-19 cases.** Where sufficient volume was available, serum from COVID-19 cases (n=184 total) from the validation set were tested using the Roche anti-nucleocapsid Elecsys antibody assay, widely used in clinical labs in the UK. Within this cohort, the sensitivity of our candidate screening assays for all COVID-19 cases, as well as target groups was considered. Confidence intervals were calculated using the Clopper method.

| | **Target group:** | **Total COVID-19** | **PCR confirmed** | **Suspected** | **Acute** | **Early Convalescent** | **Late Convalescent** |
|---|---|---|---|---|---|---|---|
| **Assay** | Number | 184 | 152 | 32 | 44 | 72 | 68 |
| **Roche N Elecsys** | Positive, n | 157 | 130 | 27 | 38 | 60 | 59 |
| | Negative, n | 27 | 22 | 5 | 6 | 12 | 9 |
| | Sensitivity, (95% CI) | 85.33 (79.37-90.05) | 85.53 (78.91-90.64) | 84.38 (67.21-94.55) | 86.36 (72.65-94.70) | 83.33 (72.70-90.95) | 86.76 (76.36-93.67) |
| **N Pan ELISA** | Positive, n | 137 | 116 | 21 | 29 | 52 | 56 |
| | Negative, n | 47 | 36 | 11 | 15 | 20 | 12 |
| | Sensitivity, (95% CI) | 74.46 (67.52-80.48) | 76.32 (68.75-82.71) | 65.63 (46.81-80.77) | 65.91 (50.08-78.99) | 72.22 (60.41-81.87) | 82.35 (71.20-90.39) |
| **RBD Pan ELISA** | Positive, n | 137 | 117 | 20 | 25 | 53 | 59 |
| | Negative, n | 47 | 35 | 12 | 19 | 19 | 9 |
| | Sensitivity, (95% CI) | 74.46 (67.52-80.48) | 76.97 (69.46-83.29) | 62.50 (43.69-78.15) | 56.82 (41.03-70.92) | 73.61 (61.90-83.06) | 86.76 (76.36-93.67) |
| **Spike Pan ELISA** | Positive, n | 164 | 141 | 23 | 34 | 65 | 65 |
| | Negative, n | 20 | 11 | 9 | 10 | 7 | 3 |
| | Sensitivity, (95% CI) | 89.13 (83.71-93.20) | 92.76 (87.42-96.31) | 71.87 (53.25-85.78) | 77.27 (62.16-88.24) | 90.28 (80.99-95.94) | 95.59 (87.64-99.07) |
| **Bridging Assay** | Positive, n | 163 | 141 | 22 | 35 | 64 | 64 |
| | Negative, n | 21 | 11 | 10 | 9 | 8 | 4 |
| | Sensitivity, (95% CI) | 88.59 (83.08-92.75) | 92.76 (87.42-96.31) | 68.75 (49.99-83.32) | 79.55 (64.70-89.96) | 88.89 (79.28-95.01) | 94.12 (85.62-98.35) |

**Table S6. Assigning international Binding Antibody Unit (BAU) values to standards and samples on in house assays.**

| | N Pan ELISA | N IgG ELISA | Spike Pan ELISA | Spike IgG ELISA |
|---|---|---|---|---|
| Standard Pool equivalent BAU/ml concentration (95% CI) | 605.15 (595.2-613.6) | 645.27 (189.03-862.51) | 439.3 (426.2-451.5) | 405.44 (329.34-469.39) |
| Parallel to WHO standard | Yes | Yes | Yes | Yes |
| F statistic (P value)[1] | 0.676 (0.4717) | 0.4461 (0.5087) | 0.0372 (0.9991) | 0.008397 (0.9771) |
| Chosen Threshold | Norm OD 0.36 | Norm OD 0.43 | 0.51 | Norm OD 0.66 |
| BAU/ml at threshold (95% CI) | 38.04 (35.11-41.06) | 74.744 (69.51-80.17) | 28.41 (26.71-30.19 | 74.69 (70.91-78.66) |

1 **-** The F statistic and P values presented here are derived from models where average ODs of each individual dilution series were considered as replicated for each standard.

**Table S7. STARD checklist.**

| Section & Topic | No | Item | Reported on page # |
|---|---|---|---|
| TITLE OR ABSTRACT | | | |
| | 1 | Identification as a study of diagnostic accuracy using at least one measure of accuracy (such as sensitivity, specificity, predictive values, or AUC) | 1 (Title page) |
| ABSTRACT | | | |
| | 2 | Structured summary of study design, methods, results, and conclusions (for specific guidance, see STARD for Abstracts) | 3 |
| INTRODUCTION | | | |
| | 3 | Scientific and clinical background, including the intended use and clinical role of the index test | 6-8 |
| | 4 | Study objectives and hypotheses | 7-8 |
| METHODS | | | |
| *Study design* | 5 | Whether data collection was planned before the index test and reference standard were performed (prospective study) or after (retrospective study) | 34-35 |
| *Participants* | 6 | Eligibility criteria | 34-35 |
| | 7 | On what basis potentially eligible participants were identified (such as symptoms, results from previous tests, inclusion in registry) | 34-35 |
| | 8 | Where and when potentially eligible participants were identified (setting, location and dates) | 34-35 |
| | 9 | Whether participants formed a consecutive, random or convenience series | 34-35 |
| *Test methods* | 10a | Index test, in sufficient detail to allow replication | 24-29 |
| | 10b | Reference standard, in sufficient detail to allow replication | 34 |
| | 11 | Rationale for choosing the reference standard (if alternatives exist) | 7-8 |
| | 12a | Definition of and rationale for test positivity cut-offs or result categories of the index test, distinguishing pre-specified from exploratory | 9-11; Table S2 |
| | 12b | Definition of and rationale for test positivity cut-offs or result categories of the reference standard, distinguishing pre-specified from exploratory | 34 |
| | 13a | Whether clinical information and reference standard results were available to the performers/readers of the index test | 32 |
| | 13b | Whether clinical information and index test results were available to the assessors of the reference standard | N/A |
| *Analysis* | 14 | Methods for estimating or comparing measures of diagnostic accuracy | 33 |
| | 15 | How indeterminate index test or reference standard results were handled | N/A |

| | | | |
|---|---|---|---|
| | 16 | How missing data on the index test and reference standard were handled | N/A |
| | 17 | Any analyses of variability in diagnostic accuracy, distinguishing pre-specified from exploratory | 33 |
| | 18 | Intended sample size and how it was determined | N/A |
| RESULTS | | | |
| *Participants* | 19 | Flow of participants, using a diagram | Figure 1 |
| | 20 | Baseline demographic and clinical characteristics of participants | Table 1, Table 3 |
| | 21a | Distribution of severity of disease in those with the target condition | Table 1 |
| | 21b | Distribution of alternative diagnoses in those without the target condition | Table 3; Table S1; Figure S4 |
| | 22 | Time interval and any clinical interventions between index test and reference standard | N/A |
| *Test results* | 23 | Cross tabulation of the index test results (or their distribution) by the results of the reference standard | Figures 2, 3. |
| | 24 | Estimates of diagnostic accuracy and their precision (such as 95% confidence intervals) | Table 2, Tables 2, S2, S4, S5. |
| | 25 | Any adverse events from performing the index test or the reference standard | N/A |
| DISCUSSION | | | |
| | 26 | Study limitations, including sources of potential bias, statistical uncertainty, and generalisability | 17-23 |
| | 27 | Implications for practice, including the intended use and clinical role of the index test | 17-23 |
| OTHER INFORMATION | | | |
| | 28 | Registration number and name of registry | N/A |
| | 29 | Where the full study protocol can be accessed | N/A |
| | 30 | Sources of funding and other support; role of funders | 3-4 |

#### Supplementary Figures

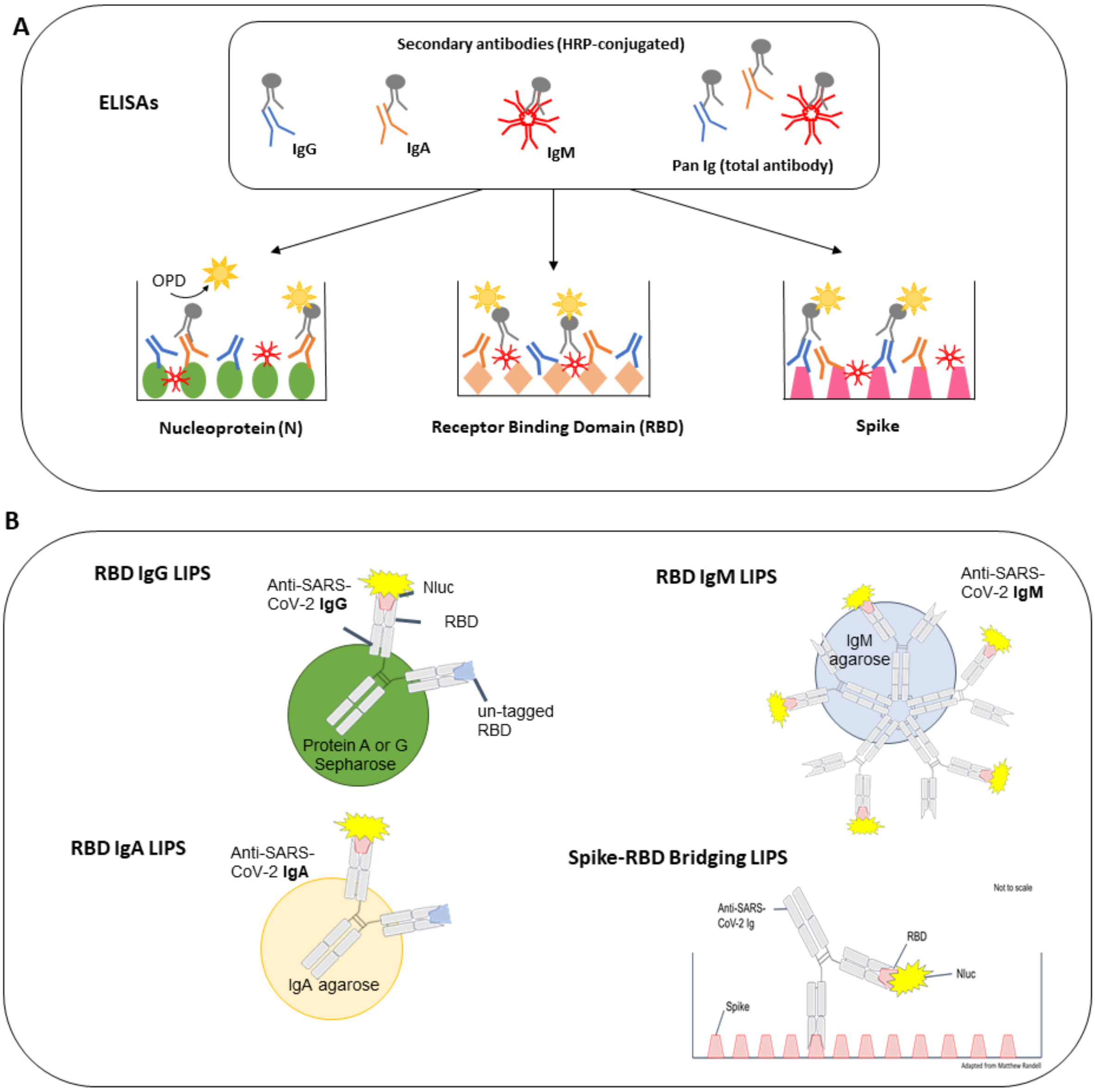

**Figure S1. Schematic of ELISA and LIPS based assays developed as part of this study.** A) ELISAs were developed to detect antibodies specific to three antigens: nucleoprotein (N), receptor binding domain (RBD) of Spike protein, and a full-length trimeric Spike. For each antigen was coated on a ELISA plates overnight prior to blocking and addition of samples. During sample incubation, specific antibodies of all isotypes (IgG, IgA and IgM) contained within the samples will bind to target antigens. Detection of antibodies is achieved by addition of one of 4 x HRP-conjugated secondary antibodies specific to either IgG, IgA or IgM (each specific to the heavy chain which determines antibody isotype) or a Fab-specific antibody which recognised total antibody (or Pan-immunglobulins (Pan-Ig)). B) The isotype specific LIPS assays are liquid phase assays which using a luciferase (Nluc) tagged RBD antigen to detected antibodies. To determine specific isotypes of antibody, the samples are first purified for IgG, IgA or IgM using specific agarose beads (Protein A or G Sepharose for IgG, IgA agarose for IgA and IgM agarose for IgM) prior to mixing with Nluc-labelled RBD. For the IgG and IgA assays, unlabelled RBD was also included (competition) as a comparator to using only labelled

RBD, to improve assay specificity (the difference between the signals is reported). To measure total antibody, a novel Spike-RBD bridging assay was developed, whereby Spike was coated onto an ELISA plate, followed by addition of sample and detection using Nluc-labelled RBD.

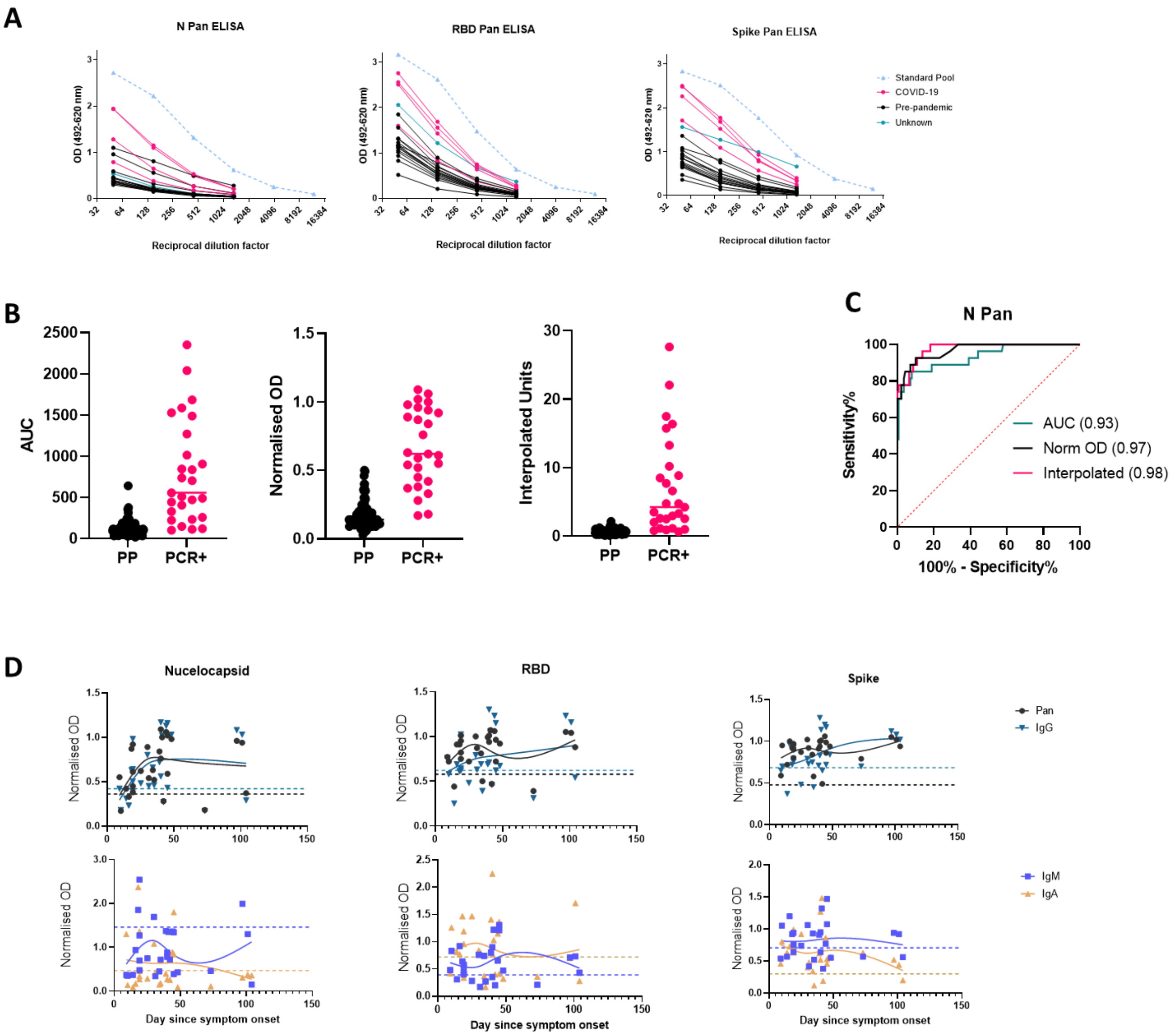

**Figure S2. Optimisation of single dilution, low-volume ELISAs and LIPS assays for detecting SARS-CoV-2 specific antibodies using a subset of the threshold set.** ELISA were developed to detect either all isotypes (Pan), or IgG, IgA and IgM antibodies specific to either Nucleocapsid (N) the receptor binding domain (RBD) of the Spike protein, or the full trimeric Spike protein for maximum discrimination between COVID-19 samples and pre-pandemic controls. A) Initially, OD adjusted for background was reported. Example plots from the N, RBD and Spike Pan ELISAs showing sample dilution series compared to a 6-point 3-fold dilution series of the pooled internal standard. B) We explored multiple ways or expressing the results from test samples, including reporting area under the curve from 4-fold dilution series, a normalised OD (sample OD normalised to top standard) and interpolated units. Data from an example assay (N Pan ELISA) is shown for the optimisation set of n=160 samples (n=27 COVID-19 cases and n=133 pre-pandemic) from the threshold set. C) Normalised OD values for n=27 samples from COVID-19 cases were compared to pre-pandemic levels (presumed non specific/background responses) for all 12 ELISA assays. Plots display Normalised OD readings for Pan (black circles) and IgG (teal triangles) assays (top panels) and IgM (blue squares) and IgA assays (orange triangle) (bottom panels). A spline (LOESS) curve was fitted to each assay data for the COVID-19 cases to indicate average trend in response over time since symptom onset. dashed horizontal lines indicate the 98the percentile of the n=133 pre-pandemic samples (i.e. the threshold to achieve 98% specificity with these samples).

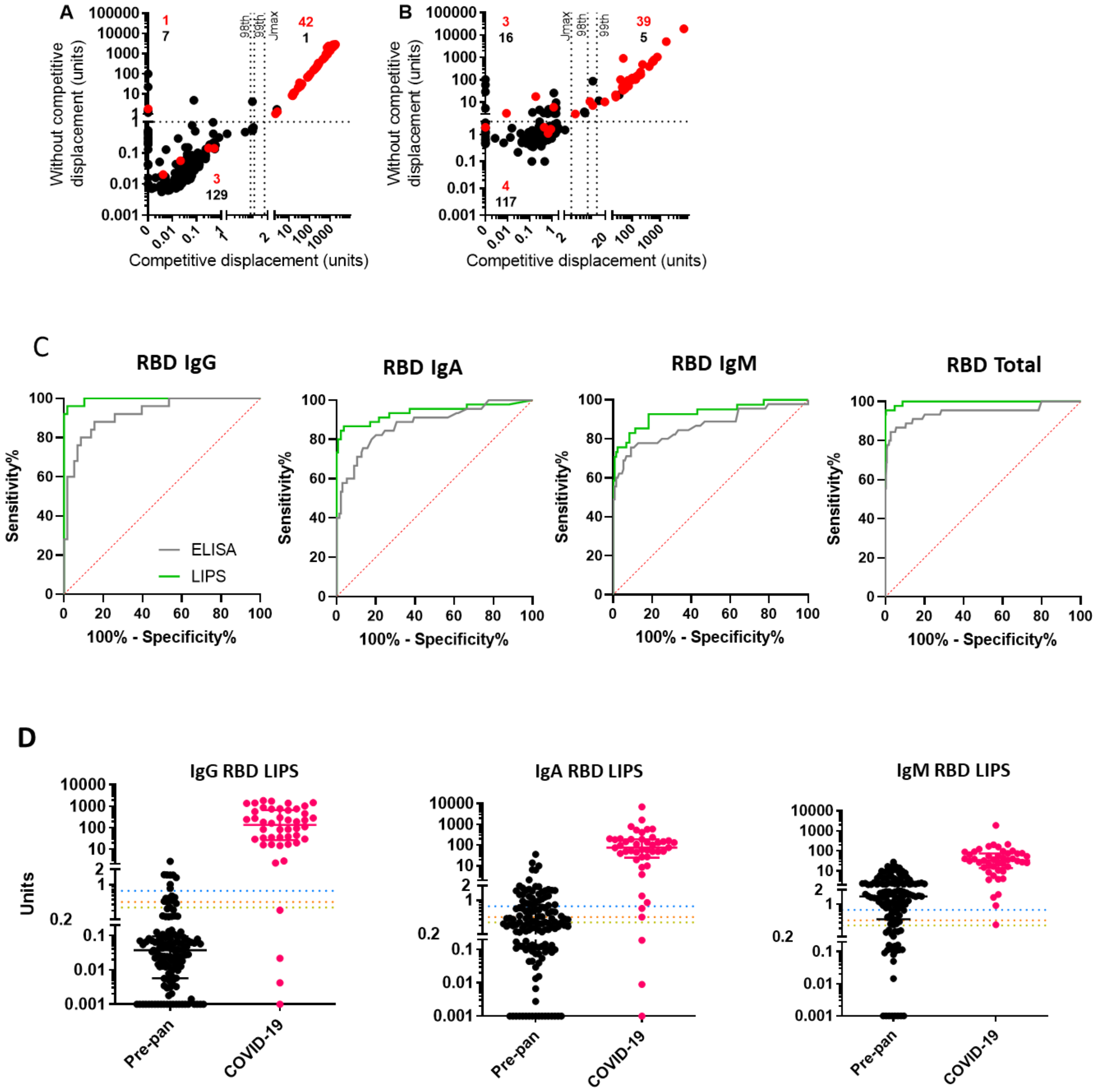

**Figure S3. Development of LIPS assays for detection of RBD specific antibodies.** RBD labelled with Nluc was used to develop assays to detect RBD-specific IgG, IgA, IgM and total antibody responses after COVID-19 infection. Competitive displacement was found to improve discrimination. For IgG LIPS (A) and IgA LIPs (B) with and without competitive displacement with un-tagged RBD (8x10$^{-8}$ mols/L). Red points and text for COVID-19 cases and black dots and text for pre-pandemic samples. Dotted lines for the x-axis represent thresholds derived from these data, for the y-axis the dotted line is a threshold that would identify all COVID-19 cases found positive by competition. C) ROC curves for ELISA and LIPS RBD-specific antibody assays in a cohort of n=132 pre-pandemic and n=45 samples from COVID-19 cases from the threshold set. D) Dot plots showing results of IgG (with competition), IgA (with competition) and IgM-specific specific LIPS RBD assays in pre-pandemic (n=58, n134, n=132 respectively) and COVID-19 cases (n=25, n=45, n=45 respectively) from the threshold set. In each plot, the candidate thresholds also displayed (1 – the 99$^{th}$ percentile of pre-pandemic levels (orange dashed line); 2 - The 98th percentile (yellow dashed line); 3 – Youden's index (blue dashed line)).

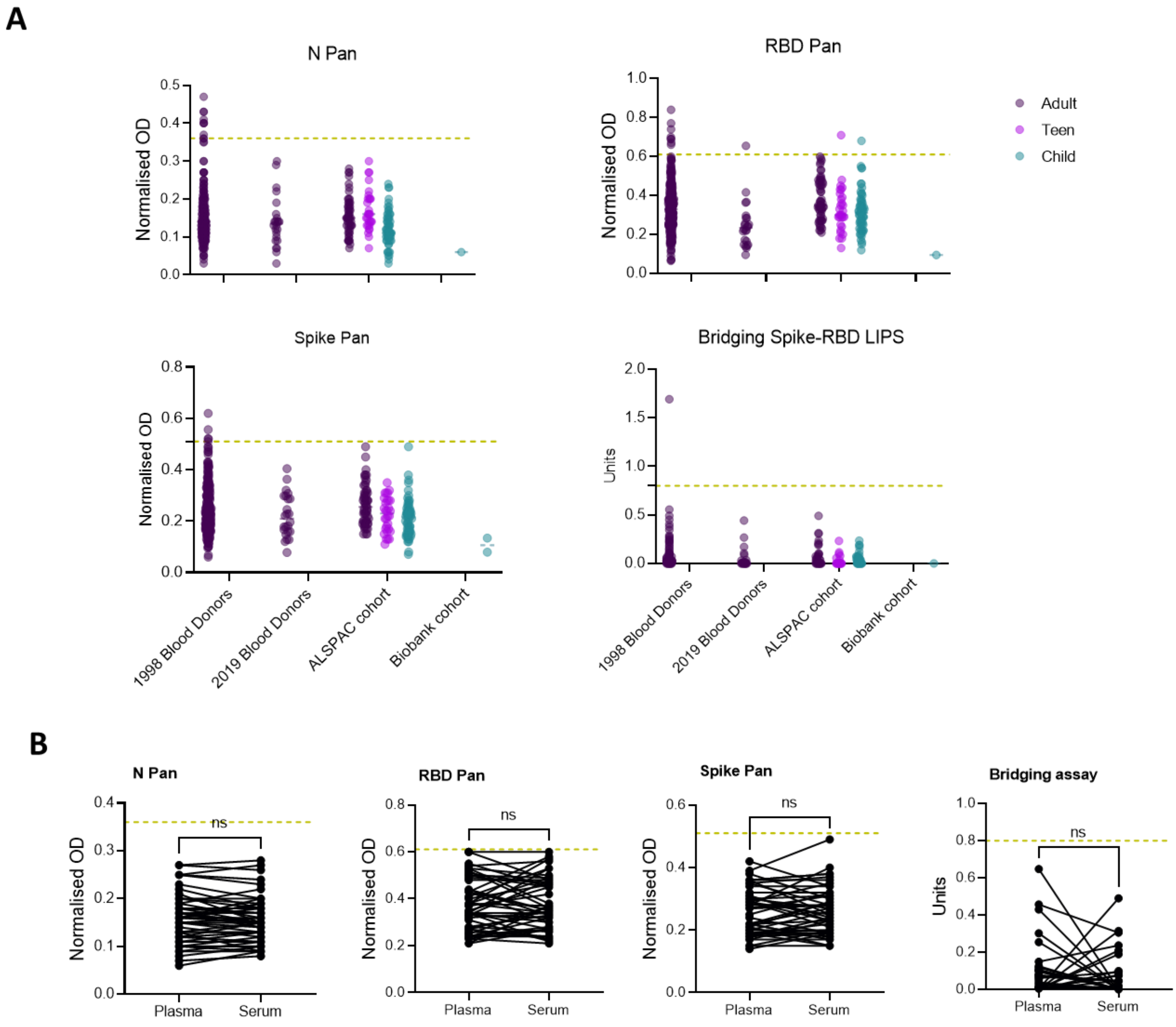

**Figure S4. Exploring factors associated with cross-reactive in known-negative pre-pandemic samples.** A) Pre-pandemic levels of total antibody from pre-pandemic samples derived from children, teens, or adults recruited as part of the ALSPAC cohort. B) Serum vs plasma results for N Pan, RBD Pan and Spike Pan ELISAs, and Spike-RBD Bridging LIPS in matched samples.

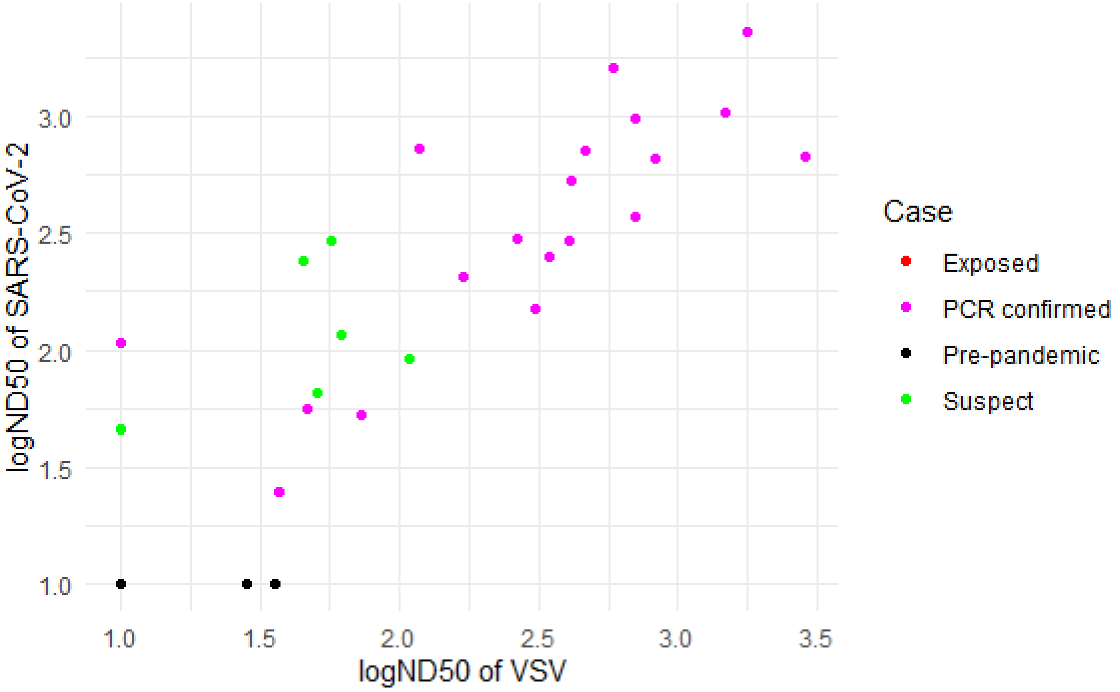

**Figure S5. Relationship between half maximal neutralisation titres measured using two different neutralisation assays.** The log10 ND50 values from the pseudotype VSV neutralisation assay (x axis) are plotted against the log10 ND50 values from the SARS-CoV-2 microneutralisation assays (y axis). Correlation was performed using the kendall's tau and the coefinnicient found to be 0.79 p value = $6.94 \times 10^{-11}$.
